# Identification of a serum proteomic biomarker panel using diagnosis specific ensemble learning and symptoms for early pancreatic cancer detection

**DOI:** 10.1101/2023.12.24.23300505

**Authors:** Alexander Ney, Nuno R. Nené, Eva Sedlak, Pilar Acedo, Oleg Blyuss, Harry J. Whitwell, Eithne Costello, Aleksandra Gentry-Maharaj, Norman R. Williams, Usha Menon, Giuseppe K. Fusai, Alexey Zaikin, Stephen P. Pereira

**Author notes:** These authors contributed equally to the work (Alexander Ney, Nuno R. Nené, Stephen P. Pereira). To whom correspondence should be addressed: Dr Alexander Ney, Dr Nuno Rocha Nené, Prof Stephen Pereira.

## Abstract

**BACKGROUND:** The grim (<10% 5-year) survival rates for pancreatic ductal adenocarcinoma (PDAC) are attributed to its complex intrinsic biology and most often late-stage detection. The overlap of symptoms with benign gastrointestinal conditions in early stage further complicates timely detection. The suboptimal diagnostic performance of carbohydrate antigen (CA) 19-9 and elevation in benign hyperbilirubinaemia undermine its reliability, leaving a notable absence of accurate diagnostic biomarkers. Using a selected patient cohort with benign pancreatic and biliary tract conditions we aimed to develop a biomarker signature capable of distinguishing patients with non-specific yet concerning clinical presentations, from those with PDAC.

**METHODS:** 539 patient serum samples collected under the Accelerated Diagnosis of neuro Endocrine and Pancreatic TumourS (ADEPTS) study (benign disease controls and PDACs) and the UK Collaborative Trial of Ovarian Cancer Screening (UKCTOCS, healthy controls) were screened using the Olink Oncology II panel, supplemented with five in-house markers. 16 specialized base-learner classifiers were stacked to select and enhance biomarker performances and robustness in blinded samples. Each base-learner was constructed through cross-validation and recursive feature elimination in a discovery set comprising approximately two thirds of the ADEPTS and UKCTOCS samples and contrasted specific diagnosis with PDAC.

**RESULTS:** The signature which was developed using diagnosis-specific ensemble learning demonstrated predictive capabilities outperforming CA19-9 and individual biomarkers in both discovery and validation sets. An AUC of 0.98 (95% CI 0.98 – 0.99) and sensitivity of 0.99 (95% CI 0.98 - 1) at 90% specificity was achieved with the ensemble method, which was significantly larger than the AUC of 0.79 (95% CI 0.66 - 0.91) and sensitivity 0.67 (95% CI 0.50 - 0.83), also at 90% specificity, for CA19- 9, in the discovery set (p=0.0016 and p=0.00050, respectively). During ensemble signature validation, an AUC of 0.95 (95% CI 0.91 – 0.99), sensitivity 0.86 (95% CI 0.68 - 1), was attained compared to an AUC of 0.80 (95% CI 0.66 – 0.93), sensitivity 0.65 (95% CI 0.48 – 0.56) at 90% specificity for CA19-9 alone (p=0.0082 and p=0.024, respectively). When validated only on the benign disease controls and PDACs collected from ADEPTS, the diagnostic-specific signature achieved an AUC of 0.96 (95% CI 0.92 – 0.99), sensitivity 0.82 (95% CI 0.64 – 0.95) at 90% specificity, which was still significantly higher than the performance for CA19-9 taken as a single predictor, AUC of 0.79 (95% CI 0.64-0.93) and sensitivity of 0.18 (95% CI 0.03 – 0.69) (p= 0.013 and p=0.0055, respectively).

**CONCLUSION:** Our ensemble modelling technique outperformed CA19-9, individual biomarkers and prevailing algorithms in distinguishing patients with non-specific but concerning symptoms from those with PDAC, with implications for improving its early detection in individuals at risk.

## Introduction

Pancreatic cancer (PC) remains lethal with approximately 500,000 new cases diagnosed globally each year with a comparable number of deaths. Pancreatic ductal adenocarcinoma (PDAC) ranks as the seventh primary cause of cancer-related mortality (1, 2). Projections suggest that by 2030, mortality rates from PDAC will exceed that of other prevalent cancers, a shift which is attributed to an increasing incidence of obesity, diabetes mellitus, alcohol consumption in some regions (Europe, North America, and Oceania) and advancements in detection and institution of screening initiatives that facilitate the timely identification of more common cancers (1-3). Across the European Union and the United Kingdom, mortality rates of PDAC have surpassed lung, breast and prostate cancers, underscoring the pressing need for enhancements in strategies for both detection and treatment of PC (4). These improvements are crucial to mitigate the growing burden of this disease (5, 6).

The overall 5-year survival for PC patients is less than 10%. These figures improve in patients diagnosed with pre-invasive lesions (intraepithelial neoplasia, mucinous cystic lesions) or small tumours (< 2cm) detected at a localised stage (7). Patients with resectable disease are only identified in less than 20% of cases and advances in early detection strategies hold potential for improving these dismal figures (8, 9). The relatively low incidence and lifetime risk for PC in the general population (1.3%) preclude asymptomatic, average-risk adult (>50 age) screening, and efforts are rather focused on high-risk populations (9-11). Internationally, screening and surveillance is therefore recommended only in high-risk individuals (genetically predisposed, family history and high-risk pancreatic cysts), where a lifetime risk of at least 5% justifies their surveillance (9, 10, 12, 13). While surveillance in these high-risk cohorts is consensus, we also reported on symptomatic cohorts in which the increased risk could justify investigations, as an additional risk group (9, 14).

Existing evidence regarding the effect of timely diagnosis on outcomes in PDAC are limited, mostly due to the lack of randomisation, appropriate statistical considerations and homogenisations of study populations, and the topic remains an area of strong debate (15). Yet with research indicating that PDAC progresses from early (T1) stage to advanced (T4) in just over a year, and larger pancreatic cancers (>2 cm) metastases are detectable within approximately 3.5 months (range between 1.2 to 8.4 months), it is very likely that prompt identification of PC would improve its prognosis (15-17).

The reality of the situation however is that disease rarity, the presence of non-localising symptoms, the relatively low positive predictive values even for cancer specific ‘red-flag’ and advanced symptoms (e.g. weight loss, painless jaundice of 4-13%) challenge timely recognition in primary care settings, and a substantial number of PC patients are diagnosed following prolonged periods of clinical uncertainty (18, 19). Previous case-control primary care studies associated various abdominal symptoms and increased frequency of primary care consultations with PDAC, over the two years preceding its diagnosis (14, 20, 21). These data suggest another potential window of opportunity for acceleration of PC detection.

In roughly 30% of patients, PC manifests in the form of jaundice indicating tumour induced biliary obstruction, which is more evident in pancreatic head tumours (22). Together with significant weight loss, these frequently represent an already advanced disease. Although most often explained by benign aetiologies, symptoms such as back or epigastric pain, dyspepsia, anorexia, bloating, changes in consistency of stool, weight loss and anxiety/depression may also indicate an underlying pancreatic malignancy (14, 20-23). Such symptoms in adults (age > 60 years) with lifestyle factors (including heavy alcohol and tobacco consumption, obesity) and on the background of new or long-standing diabetes and chronic pancreatitis, are worrisome (9, 14, 21). In such patients, the United Kingdom National Institute for Health and Care Excellence (NICE) recommends direct access to cross-sectional imaging by CT, FDG-PET/CT, or EUS within two weeks (24). Although shorter diagnostic intervals are associated with extended survival, the non-specific clinical presentation and the complexity of diagnostic pathways result in delayed referral to specialised centres (14).

To accelerate and improve cancer detection rates in the UK, ‘electronic cancer decision support tools’ (eCDST) have been developed to support primary care clinicians in fast tracking investigations in cases of suspected cancer (25-27). Risk prediction models/algorithms such as QCancer (25-27) combine symptoms data, patient risk factors and laboratory tests to predict a risk of undiagnosed cancers of various anatomical sites (colon, pancreas, renal, gastro-oesophageal and ovarian). These are digitally available for primary care physicians through patient record and data management portals (such as EMIS Web and INPS) and where higher risk justifies further investigations, could be combined with blood biomarker panels for further risk stratification prior to more invasive workup.

When suspected, establishing a diagnosis will involve measurement of the serum marker CA19-9, cross-sectional (computed tomography or magnetic resonance) imaging and histopathology (endoscopic ultrasound guided tissue biopsy; EUS-FNB). CA19-9 is most reliable as a marker of tumour resectability, prognosis and monitoring of disease progression (28, 29), but as a diagnostic marker it performs poorly (median sensitivity and specificity of ∼80%; AUC= 0.82), particularly in stage I/II disease and in Lewis body negative patients (30, 31). The development of reliable and accurate diagnostic biomarkers is essential for risk stratification and prioritisation of further investigations, as well as justification of invasive interventions where the findings on imaging are unequivocal (32).

Using serum samples collected from a selected study cohort with benign pancreatic and biliary tract conditions and applying robust machine learning stacked modelling, we therefore developed a serum biomarker signature capable of differentiating PC patients from healthy individuals and patients with benign abdominal conditions presenting with non-specific yet concerning symptoms for pancreatic cancer, at higher rates than CA19-9 and other state-of-the-art biomarkers.

## Materials and Methods

### Study Design

As our cohort, we used serum samples from the Accelerated Diagnosis of neuro Endocrine and Pancreatic TumourS (ADEPTS) study (33) (UCL/UCLH Research Ethics Committee reference 06/Q0512/106, IRAS Number 234637, NIHR portfolio no. 7343) study - an early detection study aimed at detecting pancreatic cancer in patients at an earlier stage. As part of the Early Diagnosis Research Alliance (EDRA), the ADEPTS study (previously referred to as TRANSlational research in BILiary tract and pancreatic diseases (*TRANSBIL)* study), commenced in 2018 and included a multicentre prospective blood sample collection from patients with non-specific but concerning symptoms associated with PDAC. Patients were recruited at gastroenterology/hepatobiliary and surgical clinics at University College London (UCLH) and the Royal Free Hospitals (RFH), London, UK. Blood samples were collected from subjects with benign hepatobiliary conditions as well as those with PDAC (stages I-IV). All patients recruited to the ADEPTS study provided written informed consent.

For PDAC patients, tumour staging was performed according to the AJCC 8^th^ edition (TNM) based on cross-sectional imaging and for those undergoing surgery, based on multi-disciplinary team recordings. All included PDAC cases were histologically confirmed by UCLH and RFH local pathologists based on tissue analysis obtained by endoscopic ultrasound guided fine needle biopsies or specimens obtained during surgical resection.

For benign disease controls, patients were selected to include the following diagnoses: chronic pancreatitis, intraductal papillary mucinous neoplasms (IPMN), or benign pancreatic diseases (e.g., serous cystadenomas and pancreatic heterotopia). Patients with acute and chronic pancreatitis, pancreatic cysts, benign biliary duct diseases (e.g., IgG4 disease), liver disease, gastritis/reflux disease, gallstones as well as those with familial history of pancreatic cancer, were also used. Samples also included those collected from patients presenting with non-specific symptoms which were not otherwise explained by an underlying gastrointestinal pathology (such as non-specific abdominal pain and irritable bowel syndrome) as well as other malignancies. Medical history and confirmation of diagnosis was obtained from hospital medical records and included GP and secondary clinic referral letters. For 45 patients, a QCancer score was available at time of specialist centre consultations. QCancer calculates the probability of an individual as harbouring an existing, yet undiagnosed cancer, by considering their specific risk factors and presenting symptoms. These are digitally available for primary care physicians through patient record and data management portals such as EMIS Web and INPS and designed as clinical decision support tools to aid in assessment of need for specialist referrals (34).

To further represent the healthy population we also used samples from 72 healthy control UKCTOCS (35) samples that were collected from a nested case control discovery study part of UKCTOCS reported before (36), which had been previously approved by the Joint UCL/UCLH Research Ethics Committee A (Ref. 05/Q0505/57). Written informed consent for the use of samples in the UKCTOCS trial and secondary ethically approved studies was obtained from donors and no data allowing identification of patients was provided. The original UKCTOCS dataset from which data was used here was derived from serum samples collected from post-menopausal women, aged between 50 and 74 years, who were recruited between the years 2001 and 2005 (35). The collection of these samples was conducted in accordance with a specific Standard Operating Procedure (SOP) (37, 38). For the current work our interest lies only with the UKCTOCS matched non-cancer controls, i.e., with no cancer registry code, from individual women selected based on collection date, age, and centre to minimize variation due to handling and storage. Comprehensive information regarding diabetes status for the selected UKCTOCS participants was either unavailable or incomplete. In addition, data on disease duration was not accessible. Consequently, it was not feasible to stratify samples to discovery and validation sets based on the type of diabetes they may have had. For the purposes of this study, only healthy controls that were matched to PDAC cases, with less than one year to diagnosis, were utilized.

A total of 539 serum samples (493 controls and 46 PDAC cases, see Table 1) were analysed using the Olink multiplex immunoassay Oncology II panel in addition to five in-house markers: Carbohydrate antigen 19-9 (CA19-9), Interleukin 6 Cytokine Family Signal Transducer (IL6ST/IL6RB), von Willebrand factor (VWF), Pyruvate kinase isozymes M1/M2 (PKM/PKM2) and Thrombospondin 2 (THBS2/TSP2). The selection of additional markers, beyond CA19-9, was informed by our preceding research in early detection of PDAC (36, 39). In those studies, a panel of markers was identified due to its demonstrated ability to facilitate the early detection of pancreatic cancer, with a lead time of up to two years prior to diagnosis.

**Table 1.** Cohort characteristics. The data set used to develop and test the classifiers is a combination of samples collected from ADEPTS cohort and selected controls from the UKCTOCS cohort. BMI: Body Mass Index. See Study Design in Materials and Methods section for additional details. Odds ratio (OR) and respective 95% confidence intervals are also provided in the p value column.

| Variable | Cases | Controls | p value |
| --- | --- | --- | --- |
| ADEPTS |  |  |  |
| No. samples | 46 | 421 |  |
| Stage I | 4 | - | - |
| Stage II | 15 | - |  |
| Stage III | 10 | - |  |
| Stage IV | 16 | - |  |
| Unknown | 1 | - |  |
| Mean age at sample draw (yr) (range) | 69.72 (43.00-91.00) | 57.44 (19.00-93.00) | $2.47 \times 10^{-7}$<br>OR= 1.06 (1.04 – 1.09) |
| Mean BMI (kg/m2) (range) | 24.84 (12.04-41.35) | 25.30 (15.22-39.45) | 0.47<br>OR=0.97 (0.88 – 1.06) |
| Gender |  |  |  |
| Male | 31 | 180 | 0.0015<br>OR=2.72 (1.46 – 5.27) |
| Female | 15 | 241 |  |
| Diabetes |  |  |  |
| Yes | 10<br>(8Type II, 1 Type I, 1 unspecified) | 75<br>(34 Type II, 41 unspecified) | 0.44<br>OR=1.34 (0.62 – 2.70) |
| No | 36 | 346 |  |
| Ethnicity |  |  |  |
| Caucasian | 21 | 291 | $6.56 \times 10^{-4}$<br>OR=2.02 (1.34 – 3.03) |
| Unknown | 21 | 60 |  |
| Asian | 3 | 30 |  |
| Other | 2 | 18 |  |
| Afro/Caribbean | 0 | 22 |  |
| UKCTOCS |  |  |  |
| No. samples | - | 72 | - |
| Mean age at sample draw (yr) (range) | - | 62.95 (50.44-76.86) | - |
| Mean BMI (kg/m2) (range) | - | 26.53 (17.91-42.19) | - |
| Gender |  |  |  |
| Male | - | - | - |
| Female | - | 72 |  |
| Diabetes |  |  |  |
| Yes | - | 3<br>(3 Type II) | - |
| No | - | 69 |  |
| Ethnicity |  |  |  |
| Unknown | 0 | 72 | - |

### Serum analyte measurements

All ADEPTS (33) samples were randomized for testing. Supplementary Table 1 summarizes dilution factors and coefficients of variation. CA19-9 was measured using the Mucin PC/CA19-9 ELISA Kit (Alpha Diagnostic International) according to the manufacturer, using a 1:4 serum dilution. For VWF, we resorted to the Von Willebrand Factor Human ELISA Kit (abcam) at a 1:100 serum dilution. IL6ST/IL6RB by Quantikine human soluble gp130 (R&D Systems), according to manufacturer recommendations, at a 1:100 serum dilution. THBS2/TSP2 was measured using the Quantikine Human Thrombospondin-2 Immunoassay (R&D Systems) at a 1:10 serum dilution. Pyruvate kinase M2 (PKM2) was measured with an ELISA (Cloud-Clone Corp) at a 1:10 dilution.

We outsourced tests using the multiplex immunoassay Oncology II panel from Olink on all samples. This Olink panel measured known cancer antigens, growth factors, receptors, angiogenic factors, and adhesion regulators (as detailed in Supplementary Table 2). Identical assays were performed on a subset of samples derived from the UKCTOCS study (37, 38).

To bridge the normalized protein expression values from Olink between the UKCTOCS and ADEPTs datasets, we selected a representative sample set of 16 from each cohort and plated them together. Subsequently, a correction was applied to the datasets using the statistical algorithms recommended in the Olink data normalization white paper (40). This method ensured that the data from different batches and studies were comparable, thereby enhancing the robustness and validity of the findings.

### Statistical analysis

The selected set of ADEPTS samples used in this work was partitioned into two distinct sets: a discovery subset, comprising two-thirds of the total sample size, and a validation subset, encompassing the remaining one-third. Allocation into each set was performed by stratifying for specific age ranges, diabetes status, PDAC status and control diagnosis class. For the PDAC cases, tumour stage was also used. The age stratification ranges were the following: 18<Age≤28; 29<Age≤38; 39<Age≤48; 49<Age≤58; 59<Age≤68; 69<Age≤78; Age≥79. The samples assigned to the control class were made of benign conditions such as: Sphincter of Oddi dysfunction, Pancreatic Cyst, Other Cancer, Other Biliary Duct Disease, No Relevant Diagnosis, Liver Disease, Irritable Bowel Syndrome, IgG4 Disease, Gastritis/Reflux Disease, Gallstone Disease, Familial Pancreatic Cancer, Chronic Pancreatitis, Acute Pancreatitis, Isolated LFT Derangement and Non-specific Abdominal Pain. We also added an additional set of healthy control samples collected from a nested study done in UKCTOCS samples used in a previous paper (41). The controls matched by age to the PDAC cases in the UKCTOCS cohort that had a time to diagnosis below up to one year were selected. The allocation of these controls to the discovery or validation sets was done according to the division used in our previous work (41). The number of controls and cases collected for this study can be visualized in Figure 1. UKCTOCS controls are identified as ‘Healthy’. The discovery-validation split puts the prevalence of PDAC in the discovery set at close to 8%. The prevalence of PDAC in the resulting validation was approximately 14%.

**Figure 1.**
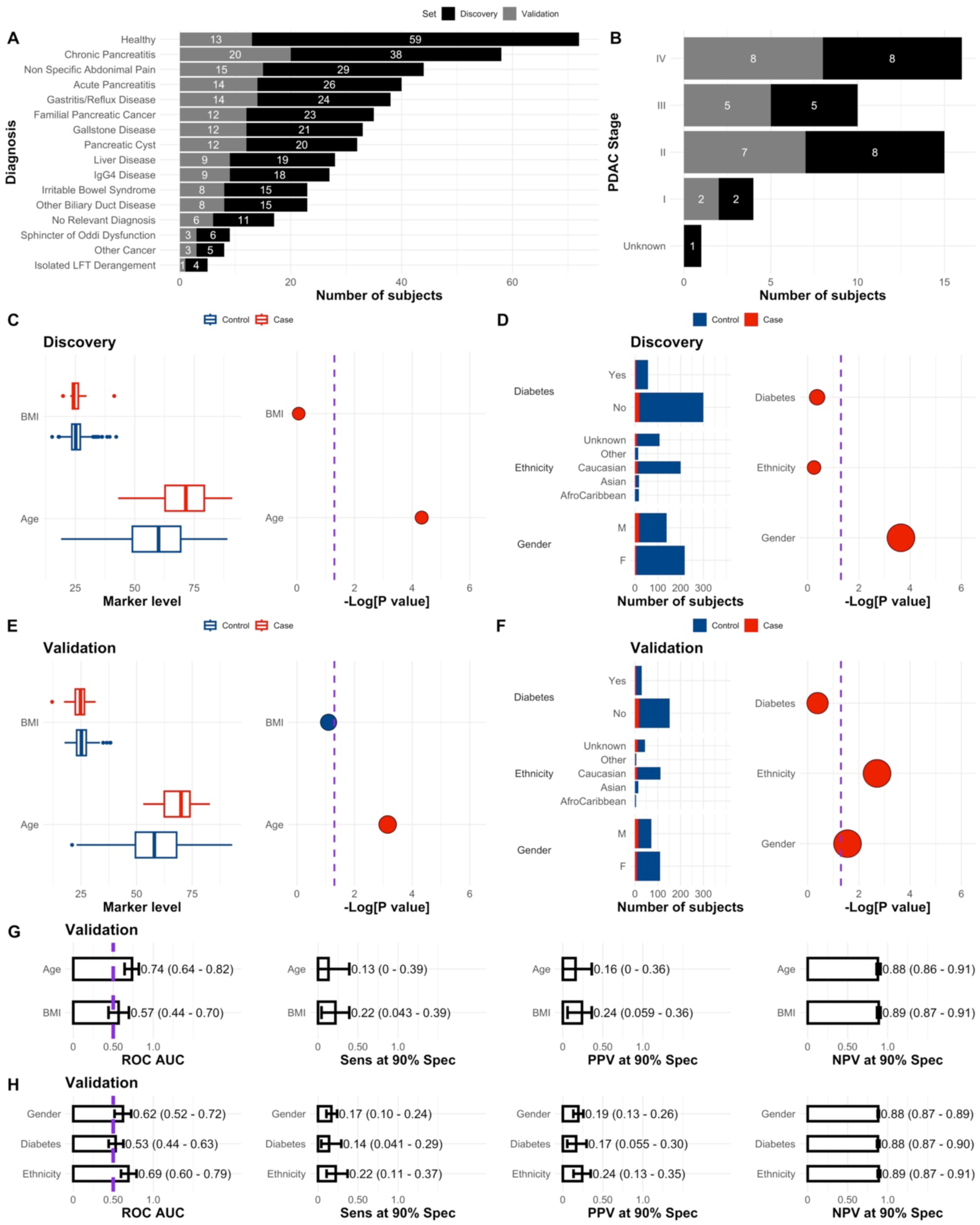
Characteristics of the discovery and validation sets. Number of controls across the discovery and validation sets **(A),** number of PDAC cases per stage **(B),** and association of BMI, Age, Diabetes, Ethnicity and Gender with PDAC status **(C-F).** In C, D, E and F dot sizes correspond to odds ratios and are colour coded according to their respective values, i.e., blue if OR<1 and red if OR>1. p values were calculated according to a logistic regression model with a bias reduction method. Purple dashed lines correspond to -Log[0.05]. **G** Receiver Operating Curve (ROC) Area Under the Curve (AUC), Sensitivity (Sens), Positive Predictive Value (PPV) and Negative Predictive Value (NPV) at 90% Specificity (Spec) performance of single marker models, i.e. BMI and Age, in the validation set. **H** Similar to A but for Gender, Ethnicity and Diabetes. Performances were calculated with the respective single feature models developed in the discovery set. The ROC AUC significance threshold is also represented by a purple dashed line at 0.5. Error bars in figures corresponding to the validation set are the 95% Confidence Intervals (CI), calculated by stratified bootstrapping 2000 times. See Statistical Analysis in Methods (main text) for further details and Supplementary Table 6, 7 and 8.

Receiver operating characteristic (ROC) curves were constructed for each model to assess diagnostic performance. The area under the curve (AUC) for the ROC curves was used as the metric. Models and techniques were compared based on their rank in the discovery under a 10-time repeated 5-fold cross-validation resampling strategy. ROC curves were generated with the *pROC* R package (version 1.18.0, https://cran.r-project.org/web/packages/pROC/index.html). 95% CI for AUCs were determined by stratified bootstrapping. All AUC confidence intervals crossing 0.5 were considered to be non-significant. P values comparing ROC curves were also calculated using the *pROC* package, under a one-sided bootstrap approach with 10000 runs.

In order to evaluate the association between each of the single markers available for this work, including clinical covariates (see Table 1 and Figure 1), and PDAC status, we used a logistic regression model implemented in the *logistf* R package (https://cran.r-project.org/web/packages/logistf/index.html, version 1.24.1). This approach fits a logistic regression model using Firth’s bias reduction method. The reported confidence intervals for odds ratios and tests were based on the profile penalized log likelihood and incorporate the ability to perform tests where contingency tables are asymmetric or contain zeros. The performance of single marker models was also verified in the discovery and validation sets (see Supplementary Figure 1 and Supplementary Table 3 and 4). The same package was also used to verify the association of the presence of symptoms and PDAC status (see Figure 4).

A comprehensive multi-dimensional examination of the collated data was conducted by employing two distinct analytical frameworks. The first was a stacked ensemble algorithm where base-learners were developed according to the same algorithm but in subsets of the discovery set where samples belonging to a specific control diagnosis class were contrasted against the same 24 PDAC cases (see for example the proportions in Figure 1). The resulting base-learners were then stacked by a logistic regression model, (see Supplementary Table 5 for the resulting coefficients and Supplementary Figure 2 for the stacking procedures). This approach aimed to leverage the predictive power of multiple models, thereby enhancing the robustness and potentially leading to more precise predictive outcomes (36, 42, 43). For each base-learner classifier we resorted to a Recursive Feature Elimination (RFE) routine with logistic regression as the fitting algorithm available through *caret* (version 6.0-93, https://cran.r-project.org/web/packages/caret/index.html) and oversampling of the minority class. This secures robust selection of features when combined with cross validation and the selection process is encapsulated inside an outer layer of resampling (44-46). Due to the prevalence of PDAC cases in the whole dataset being low, random under sampling of the majority class, here benign and healthy controls, would not have been sufficient to meet the demands of most algorithms. Therefore, creating an ensemble of classifiers specialised in contrasting a specific diagnostic class against PDAC allowed us to create more balanced subsets leading to increased performance (Figure 2). For the samples collected from UKCTOCS no symptoms information was available and, therefore, we created a separate classifier associated with this subset of individuals.

**Figure 2.**
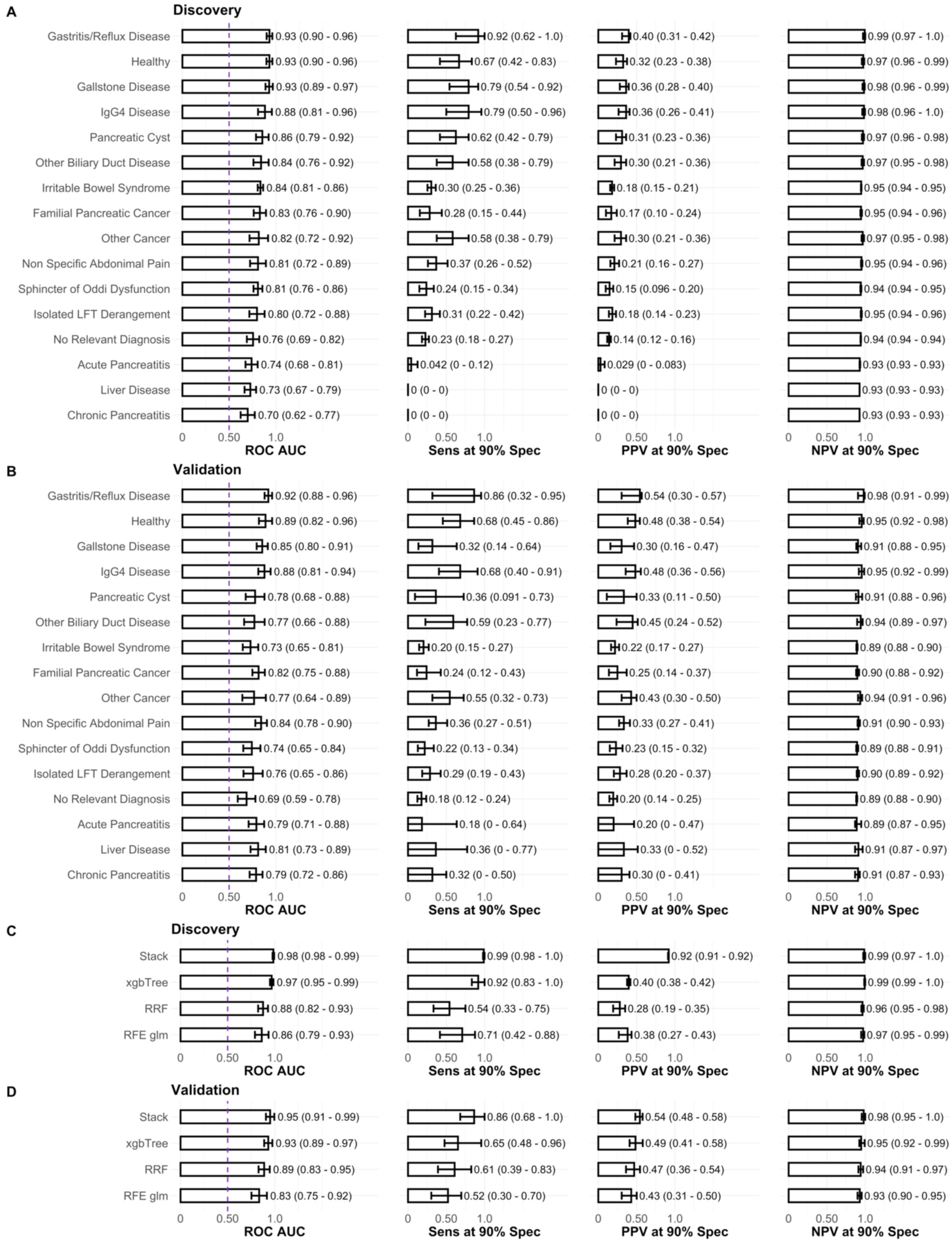
Performance of individual base-learner classifiers, stack ensemble and state-of-the art algorithms. **A** Base-learners performance in the discovery set. Each base-learner classifier was developed by training with a recursive feature elimination technique (RFE) and logistic regression (glm) in samples belonging to each specific diagnosis class against the same 24 PDACs in the discovery set. The performance reported in A is, nevertheless, of each classifier in the whole discovery set. The performances reported in **B** correspond to the base-learners developed in the discovery set but applied to the whole validation set. In **C** and **D** the performance of the ensemble stack based on the base-learners presented in A and B, as well as of state-of-the-art algorithms (xgbTree, RRF and RFE glm) is reported in the discovery and validation sets, respectively. xgbTree, RRF and RFE glm were trained in the whole discovery set, which contrasts with the ensemble algorithm.

In addition to the stacked approach we also fitted state-of-the-art algorithms such regularized random forests (RRF, version 1.9.4, https://cran.r-project.org/web/packages/RRF/index.html); extreme gradient boosting trees (xgbTree, version 1.6.0.1, https://cran.r-project.org/web/packages/xgboost/index.html); and a generalized linear model with RFE applied to the whole discovery set (RFE glm, in *caret*). The latter allowed for testing if the division into diagnosis specific classifier ensembles was advantageous at low but representative prevalence (Figure 2) in a situation where we want to contrast PDAC cases with confounding diseases in a clinical setting. All base-models were trained by 10 times repeated 5-fold cross-validation with over-sampling of the minority class (see Table 1 and Supplementary Tables 6 and 7 for information on prevalence).

To verify if the PDAC index developed with the ensemble stacked approach had any association with metrics used in the clinic but not taken into account in any stage of algorithm training, we also gathered the QCancer score (47) for individuals in the ADEPTS study (see Figure 6).

The procedure for assessing feature importance in each base learner was a model-agnostic method based on a simple feature importance ranking measure (48), implemented in the R package *vip* (version 0.3.2, https://cran.r-project.org/web/packages/vip/index.html). The model-agnostic interpretability, by decoupling the interpretation from the model itself, introduces a level of flexibility that enables its application across any supervised learning algorithm. Despite the algorithm used for each diagnosis-specific classifier being the same, the model-agnostic approach allows us to be able to generalise the computed importances to other work in the literature.

Enrichment analysis for each of the signatures developed was performed with the *gprofiler2* R package (version 0.2.1, https://cran.r-project.org/web/packages/gprofiler2/index.html). A threshold for multiple comparison correction under the framework of false discovery rate was instituted at 0.05.

## Results

### Data set characteristics

In the full set of samples collected from the ADEPTS cohort, age at the time of sample collection, 57.44 (range from 19.00 to 93.00) for controls and 69.72 (range from 43.00 to 91.00) for PDAC cases, emerged as a risk factor (OR= 1.06 (95% CI 1.04 – 1.09), p=2.47×10^-7^) (Table 1). As a predictor in a logistic regression model age as a feature achieved a ROC AUC of 0.73 (95% CI 0.66-0.79), with a cut-off at 61.5 years (calculated using the Youden J statistic). This finding was also observed in both the discovery (Supplementary Table 6 and Figure 1, ROC AUC 0.74 (95% CI 0.64-0.83), cut-off at 70) and validation sets (Supplementary Table 7 and Figure 1, 0.74 (95% CI 0.64-0.82), cut-off at 60), which incorporated not only ADEPTS samples but also healthy control samples collected from UKCTOCS (35). In our past research which was focused exclusively on UKCTOCS longitudinal samples, age similarly emerged as a risk factor for PDAC (36). Furthermore, gender (OR=2.72 (95% CI 1.46 – 5.27), p=0.0015) and ethnicity taken as a one-hot encoded variable (OR=2.02 (95% CI 1.34 – 3.03), p=6.56×10^-4^) were also confirmed as significantly associated with an increased risk of PDAC (Table 1). In the whole set of samples collected from the ADEPTS cohort, men had a 2.72-fold risk of PDAC compared to their female counterparts. Individuals of Caucasian ethnicity demonstrated a decreased risk of PDAC in a one versus rest calculation (OR=0.38 (95% CI 0.20 – 0.69), p=0.0018) and no significant association was found between PDAC risk and Asian or Afro-Caribbean ethnicity in the ADEPTS dataset under the same modelling framework (Table 1). The association of gender and PDAC was also confirmed in the discovery (OR=4.98 (95% CI 2.08 – 13.50), p=0.00023, Supplementary Table 6 and Figure 1) and validation sets (OR=2.65 (95% CI 1.11 – 6.58), p=0.028 Supplementary Table 7 and Figure 1), but ethnicity, taken as a one-hot encoded variable, remained a significant predictor of PDAC only in the validation set (OR=2.66 (95% CI 1.42 – 5.17), p=0.0020) (Supplementary Table 7 and Figure 1), which as was highlighted above also includes healthy control UKCTOCS samples. Within the group of the clinical covariates only age and gender are significant predictors of PDAC in both the discovery and validation set (Figure 1), with only age achieving a significant AUC in the validation set between these two. However, this was concomitant with remarkably low sensitivity (Sens), positive predictive (PPV) value and negative predictive value (NPV) at 90% specificity (Spec): AUC 0.74 (95% CI 0.64-0.82), Sens 0.13 (95% CI 0 - 0.39), PPV 0.16 (95% CI 0 - 0.36), NPV 0.88 (95% CI 0.86 – 0.91).

### Development of PDAC biomarker signature in the presence of confounding conditions

To aid the early detection of this cancer in individuals at risk, we aimed to develop a biomarker signature that could be used to differentiate between suspected PDACs and benign biliary conditions that often overlap in clinical presentation. We applied a uniquely developed ensemble learning model, with a logistic regression stacking layer (see Supplementary Figure 2 and statistical analysis in the Methods section), to a set of 539 serum samples (493 controls and 46 PDAC cases) which were analysed using the Olink Oncology II panel as well as four additional biomarkers we previously reported on (36). These included IL6ST, VWF, THBS2 and CA19-9. The oncogenic and prognostic glycolytic enzyme PKM2 was additionally selected based on our past report of its diagnostic utility in biliary tract cancer patients (49-51).

The application of stacked ensemble modelling as presented herein bolsters the robustness of predictive outcomes, enhancing the performance of biomarker panels through the incorporation of serum biomarker levels and relevant clinical covariates for distinct diagnostic classes. Each component classifier within the ensemble is designed to provide a specialized distinction between confounding diagnoses and PDAC, thereby establishing a heterogeneous set of classifiers that facilitates the precise identification of PDAC (see statistical analysis section in Methods). Previous studies have attested to the beneficial role of ensemble methods in augmenting early detection of PDAC against only healthy controls (36). The implementation of stacked (Stack, Figure 2), specialized classifiers, developed within the discovery set, generated a biomarker signature capable of predicting PDAC with an AUC of 0.98 (95% CI 0.98 – 0.99), sensitivity of 0.99 (95% CI 0.98 - 1), PPV 0.92 (95% CI 0.91 - 0.92) and NPV 0.99 (95% CI 0.97 - 1) at 90% specificity. In contrast, the predictive efficacy of CA19-9 in the discovery set taken as a single predictor under a logistic regression model was 0.79 (95% CI 0.66 – 0.91) (p=0.0016 under a one-sided bootstrap test comparing the two AUCs), sensitivity 0.67 (95% CI 0.50 - 0.83), PPV 0.32 (95% CI 0.26 - 0.38) and NPV of 0.97 (95% CI 0.96 - 0.99) at 90% specificity (see Supplementary Table 3). Amongst all biomarkers, CA19-9 demonstrated the most significant association (refer to Supplementary Table 3 and Supplementary Figure 1 for univariate trend associations across the discovery set), and one of the highest performances in the validation set (Supplementary Figure 1 and Supplementary Table 4).

In the validation set, the ensemble signature predicted PDAC with an AUC of 0.95 (95% CI 0.91 – 0.99), sensitivity 0.86 (95% CI 0.68 - 1), PPV 0.54 (95% CI 0.48 - 0.58) and NPV of 0.98 (95% CI 0.95 - 1) at 90% specificity. Once again, this is an improvement with respect to CA19-9 (p=0.0082, one-sided bootstrap test) taken as a univariate model developed in the discovery set; this CA19-9 model predicted PDAC status with an AUC of 0.80 (95% CI 0.66 – 0.93), sensitivity 0.65 (95% CI 0.48 - 0.56), PPV 0.49 (95% CI 0.41 - 0.56) and NPV of 0.95 (95% CI 0.92 - 0.98) at 90% specificity in the validation set. If we further validate only on the benign disease controls and PDACs collected from ADEPTS, the diagnostic-specific ensemble signature achieved an AUC of 0.96 (95% CI 0.92 – 0.99), sensitivity 0.82 (95% CI 0.64 – 0.95) at 90% specificity. This performance is also significantly higher than the performance of CA19-9 in a univariate model: AUC of 0.79 (95% CI 0.64-0.93) (p= 0.013 when compared with the full signature, one-sided test) and sensitivity of 0.18 (95% CI 0.03 – 0.69).

A closer examination of the individual performances of each base-learner classifier (Figure 2A and B) reveals that the logistic regression stacked ensemble approach has superior performance in both discovery and validation sets. Despite the best base-learner being trained on samples diagnosed as ‘Gastritis/Reflux Disease’ (Figure 2A and B), its performance was also superseded by the AUC computed with the stack model, the logistic regression coefficients of which are delineated in Supplementary Table 5. The stack model significantly relies on the “Healthy”, “Chronic Pancreatitis”, “IgG4 Disease”, “Irritable Bowel Syndrome”, ‘Other Biliary Duct Disease”, “Sphincter of Oddi Dysfunction”, “No Relevant Diagnosis”, “Other Cancer” and “Pancreatic Cyst” base-learners. Even though the remaining diagnostic class base-learners, including "Gastritis/Reflux Disease", did not reach statistical significance (p<0.05), employing a stack that solely resorts to significant base-learners led to a reduction in generalization capacity: AUC 0.98 (95% CI 0.97 – 0.99), sensitivity 0.98 (95% CI 0.95 –1), PPV 0.92 (95% CI 0.91 – 0.92), NPV 0.97 (95% CI 0.94 – 0.99) in the discovery set; AUC 0.93 (95% CI 0.87 – 0.99), sensitivity 0.82 (95% CI 0.64 – 0.95), PPV 0.53 (95% CI 0.47 – 0.57), NPV 0.97 (95% CI 0.95 – 0.99) in the validation set. Although the differences are not substantial, we retain the full set of base-learners to enhance the generalization capacity for predicting PDAC in unseen data sets and new samples.

The employment of stacked diagnosis-specialized classifiers surpassed the AUC performance of state-of-the-art algorithms such as random forests (RRF) and extreme gradient boosting methods (xgbTree), in terms of AUC, sensitivity, positive predictive value, and negative predictive value at 90% specificity (Figure 2C and D); although the performance AUC of the stacked classifier was only marginally significantly higher than that obtained with RRF (p=0.040, one-sided) and not significant when compared with xgbTree (p=0.26, one-sided), the sensitivity values at 90% specificity obtained with the alternative methods were, in fact, significantly lower, p=0.028 and p=0.045, respectively. The ensemble also outperformed a logistic regression model with recursive feature elimination (Figure 2C and D) that did not rely on ensemble modelling (p=0.0066, one-sided), further substantiating our choice of machine learning paradigm for facilitating the identification of PDAC cases in a clinical setting where confounding diagnoses may be present, and the prevalence is low.

The comprehensive index signature, incorporating all diagnostic categories, was constituted by 49 features, of which 44 were proteins (see Figure 3 for the importance associated with each). Among these proteins, 21 demonstrated a significant association with PDAC in the discovery set; ICOSLG, GPNMB, ESM-1, DLL1, VWF, ERBB2, FCRLB, CEACAM5, EGF, CTSV, FASLG, Creatinine, CPE, CA9/CAIX, TBIL, CD207, CRP, CDKN1A, EPHA2, ITGAV, and MUC-16 (see Supplementary Figure 1 and Supplementary Tables 3). The remaining 23 proteins, namely CXCL13, ERBB3, FOLR1/FR-alpha, FADD, ERBB4, CD27, AREG/AR, ADAM-TS-15, ABL1, ANXA1, CXCL17, CD70, CEACAM1, CD48, IL6ST, CD160, PKM/PKM2, CYR61/CCN1, CRNN, ADAM-8, FOLR3/FRgamma, THBS2, GZMB, did not demonstrate a significant association with PDAC in univariate models (see Supplementary Figure 1 and Supplementary Tables 3 and 4). Additionally, five clinical covariates—Gender, Age, Ethnicity, Diabetes, and Body Mass Index (BMI)—were identified as important predictors following comprehensive recursive feature elimination during cross-validation (Figure 3).

**Figure 3.**
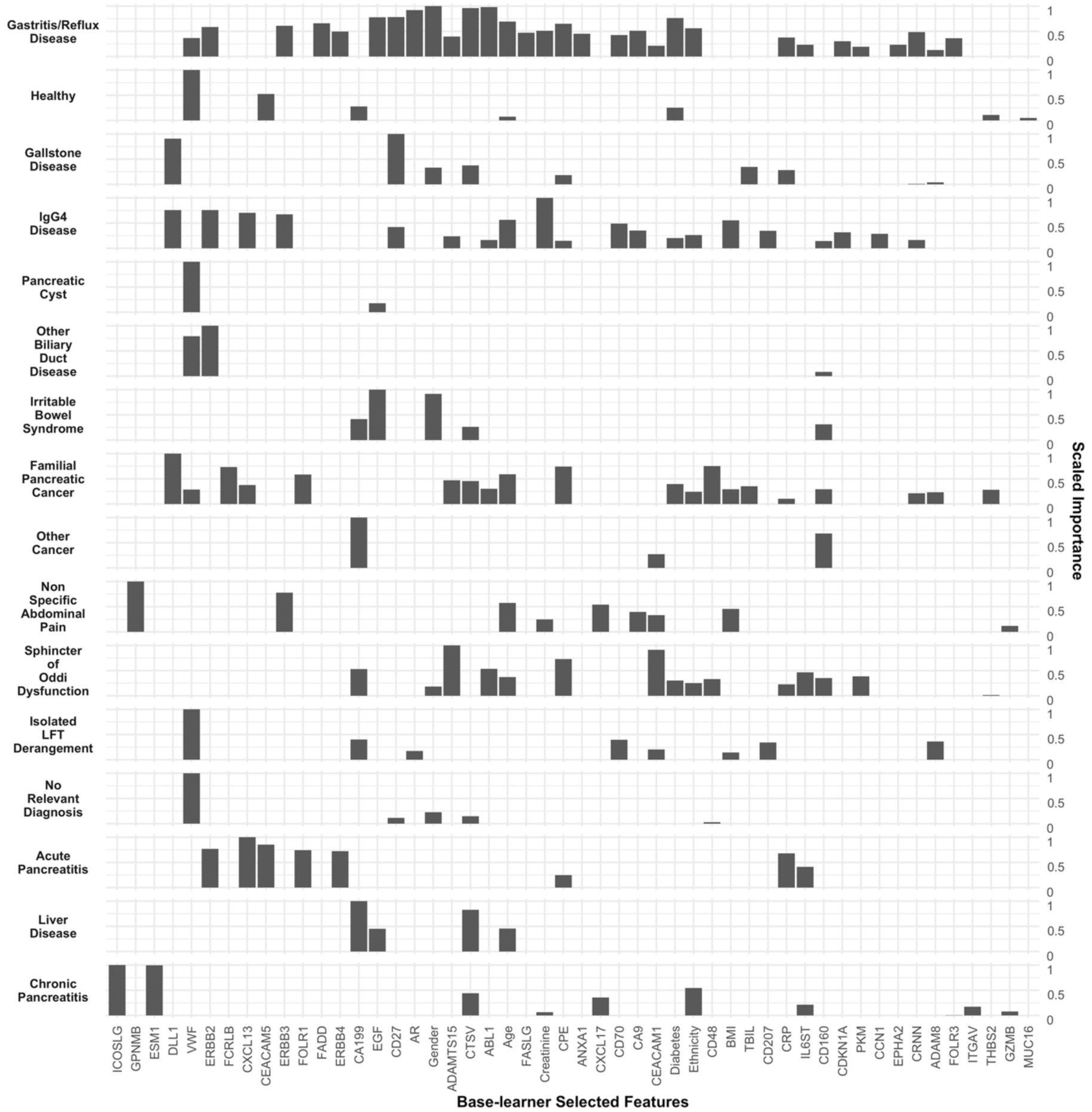
Features selected per diagnosis class (base-learner classifiers). The scaled importance is calculated within each base-learner (Figure 2A). Selected features are ranked from left to right according to the average scaled importance across base learners. See Figure 1, Supplementary Figure 1 and Supplementary Tables 3 and 4 for the univariate predictive performances of each of the markers in the discovery and validation sets. See Methods section for details on model-agnostic algorithm for feature importance calculation.

Gene Ontology (GO) and biological pathway enrichment (Kyoto Encyclopaedia of Genes and Genomes; KEGG, Reactome Pathway Database; REAC and WikiPathways; WP) analysis was performed for the selected set of features using g:Profiler (Supplementary Figure 3). Top significant terms for biological processes (BP) included ‘circulatory system development’, ‘blood vessel morphogenesis’, ‘cell adhesion’, ‘angiogenesis’, ‘blood vessel development’, ‘regulation of cell adhesion’, ‘positive regulation of cell population proliferation’, ‘cell-cell adhesion’, and ‘regulation of developmental process’. Top relevant biological pathways included: ‘PI3K-KAT signalling pathway’, ‘ERBB signalling pathway’, ‘pathways in cancer’, ‘proteoglycans in cancer’, ‘platinum drug resistance’, ‘prostate cancer’, ‘type I diabetes mellitus’, ‘MAPK signalling pathway’ and ‘focal adhesion’.

The scaled importance of each feature and diagnostic class/classifier is depicted in Figure 3. It is of significance to note that not every biomarker was selected by each individualized classifier, highlighting the requirement for an array of diverse predictors, each tailored to specific underlying conditions, to effectively identify PDAC. This is consistent with the idea that heterogeneous ensembles are fundamental for predictive capacity in blind datasets (36, 43).

Of the five selected clinical covariates, only Age, Ethnicity, and Gender manifested as significant predictors of PDAC in the validation set, as illustrated in Figure 1 and explained in the data set characteristics subsection (see also Supplementary Table 6 and 7). It is worth emphasizing that the lack of significant association between certain markers and PDAC in the discovery set does not preclude their inclusion in the signature. These variables were selected due to their contribution to the enhanced robustness and generalization capacity in predicting PDAC during cross-validation with a recursive feature elimination routine (see Methods). A similar trend was verified in prior work focussed on ensemble models for PDAC early detection against healthy controls (36).

### Application of a reduced, 8-marker signature as a differentiator of PDAC from healthy and benign controls

Across all conditions, 8 features with relatively higher scaled importance that differentiated controls from PDAC patients were selected (Figure 3). Importance is measured by the contribution of a specific feature to the output of the model (see Methods), in our case the probability of PDAC. These included CA19-9, VWF, CPE, CTSV, CEACAM1 and CD160 together with Diabetes and Age as clinicodemographic variables. Diabetes was a predictor of the differences between PDAC against familial cases, gastric reflux disease (GORD), sphincter of oddi (SOD) dysfunction, as well as healthy controls.

CA19-9 levels were only selected as a top discriminating feature against PDACs in patients with suspected sphincter of Oddi dysfunction, benign liver disease, irritable bowel syndrome (IBS), those with isolated LFT derangements as well as distinguished healthy subjects and those with other cancers (Figure 3 and Supplementary Table 8), from PDAC patients.

Von Willebrand Factor (VWF) levels differentiated PDAC from symptomatic patients with pancreatic cysts, benign biliary duct diseases, non-abdominal conditions, patients with family history of PDAC, those with GORD as well as healthy subjects.

The immunoglobulin like surface antigen molecule CD160 (peripheral natural killer cells and CD8^+^ T lymphocytes) (52) and a proposed immune checkpoint inhibitor, was selected as a significant differentiator of PDAC from benign biliary tract diseases (IgG4 disease), SOD dysfunction, IBS as well as in familial pancreatic cancer subjects and other cancers. Cathepsin V (CTSV) levels were also a predictor of multiple conditions against PDAC, including benign biliary diseases and in subjects belonging to the familial PC cohort. In healthy subjects, however, this feature did not show significant importance as a differentiator from PDAC.

Serum levels of the metallo-carboxypeptidase E (CPE) were a feature selected as significant in five conditions (acute pancreatitis, gallstones and IgG4 disease, SOD dysfunction and GORD) as well as a differentiator in those with FH of PC. A higher scaled importance was attributed to this enzyme against CA19-9 when differentiating acute and chronic pancreatitis (CP), isolated LFT derangements, unexplained abdominal pain and non-abdominal conditions versus PDAC (Figure 3).

THE CEA cell adhesion molecule (CEACAM1) was selected as a feature in patients with non-explained recurrent abdominal pain, isolated LFT derangements, GORD, SOD dysfunction as well as a feature selected against non-pancreatic cancers.

Chronic pancreatitis (CP) is a known risk factor for PDAC. In our index signature, the protein markers selected against PDAC included ESM1, ICOSLG, CTSV, CXL17 (CXC motif chemokine ligand 17), IL6ST, ITGAV, GZMB (granzyme B; secreted serine protease), with a reduced risk for cancer in Caucasian ethnicity (53-55).

We therefore opted to assess their combined performance against CA19-9 as a single marker. Using a similar stacking procedure as before, a reduced model was trained using the same ensemble approach as that highlighted before but with only 8 features as the input. The reduced signature predicted PDAC still with a high AUC value of 0.97 (95% CI 0.95-0.98), sensitivity 0.98 (95% CI 0.95-1), PPV 0.92 (95% CI 0.91-0.92) and NPV of 0.98 (95% CI 0.94-1) at 90% specificity, in the discovery set (Supplementary Figure 4C). In the validation set, however, the performance of the 8-marker signature was significantly reduced (p= 0.00038, one-sided) compared to the full stacked model (AUC of 0.84 (95% CI 0.75-0.94), sensitivity 0.64 (95% CI 0.36-0.82), PPV 0.47 (95% CI 0.33-0.53) yet with a NPV of 0.95 (95% CI 0.91-0.97) at 90% specificity (Supplementary Figure 4D)), and only marginally superior to CA19-9 as a single marker (p=0.18, one-sided). On the other hand, the 8-marker signature still outperformed CA19-9 by a relatively large margin when predicting PDAC against healthy UKCTOCS controls in the validation set: AUC_redsig_ of 0.93 (95% CI 0.84 - 1), sensitivity_redsig_ of 0.86 (95% CI 0.54 - 1), PPV_redsig_ 0.94 (95% CI 0.90 – 0.94), NPV_redsig_ 0.80 (95% CI 0.54 – 1); AUC_CA19-9_ of 0.84 (95% CI 0.70 - 0.97), sensitivity _CA19-9_ of 0.68 (95% CI 0.5 – 0.91), PPV _CA19-9_ 0.92 (95% CI 0.89 – 0.94), NPV _CA19-9_ 0.62 (95% CI 0.52 – 0.85), at 90% specificity. Under a bootstrap test this AUC difference is significant p= 0.025 (one-sided). In addition, it also outperformed the full PDAC ensemble model when predicting PDAC against healthy controls in the validation set, although the differences were not significant (p=0.2, one-sided): AUC_sig_ of 0.90 (95% CI 0.77 – 1), sensitivity_sig_ of 0.86 (95% CI 0.54 – 1), PPV_sig_ 0.94 (95% CI 0.92 – 0.94), NPV_sig_ 0.80 (95% CI 0.66 – 1) at 90% specificity.

If on the other hand the reduced signature is validated in ADEPTS samples only, i.e. in PDACs plus benign disease controls, the performance of the reduced signature is far inferior to the full signature: AUC_redsig_ of 0.83 (95% CI 0.73 – 0.93) (p= 0.0009 when compared with AUC_sig_, one-sided test), sensitivity_redsig_ 0.59 (95% CI 0.27 – 0.82), PPV_redsig_ 0.47 (95% CI 0.29 – 0.55) and NPV_redsig_ 0.94 (95% CI 0.89 – 0.97), at 90% specificity. This further justifies the use of the full ensemble signature in blind data sets and in a scenario where there is limited information on a patient trajectory, despite its increased complexity.

In consideration of the marker importance in the reduced model, no marker received a null significance across all diagnostic specific base-learners, a divergence from observations in the comprehensive signature. CA19-9 emerged with the largest average importance across conditions, which also contrasted with the full model. Age and VWF were ranked with elevated average significance across diverse conditions. It’s pertinent to note that CPE levels manifested diminished scaled importance in discerning healthy controls from PDAC, particularly when juxtaposed against CA19-9, age, and CEACAM1 (refer to Supplementary Figure 5).

### Application of the full PDAC ensemble signature in symptomatic patients

Our subsequent aim was to explore whether specific clinical manifestations were correlated with PDAC status in our ADEPTS patient cohort, for which such information was available (refer to Supplementary Figure 6 and Supplementary Table 9). As a similar type of data was not available for the UKCTOCS subset (healthy controls) used in this work, we focussed this section on the ADEPTS cohort.

In our prior research, we analysed 12 "red-flag" symptoms reported by patients up to 22 months before the diagnosis of pancreatic cancer was established (20). In this work, ‘Vomiting’ (p=0.17), ‘Asymptomatic LFT Derangement’ (p=0.28), ‘Back pain’ (p=0.54), ‘Change in Bowel Habit’ (p=0.67) and ‘Rectal Bleeding’ (p=0.76) were selected for PDAC (versus benign disease controls), yet only ‘Jaundice’ (p=3.22×10^-15^), and ‘Weight Loss’ (p=1.44×10^-6^) were significantly associated with PC cancer cases in the set of samples randomly selected from the ADEPTS cohort, in which the biomarker panel was tested (Figure 4B and Supplementary Table 9). Unsurprisingly, ‘Reflux’ (p=0.022) and ‘Bloating’ (p=0.048) were significantly associated with benign controls. Interestingly, ‘Abdominal Pain’, ‘Heartburn’, ‘Anaemia’, and ‘Dysphagia’ upon presentation were aligned more with the benign control cohort, albeit not significantly (refer to Figure 4 and Supplementary Table 9).

**Figure 4.**
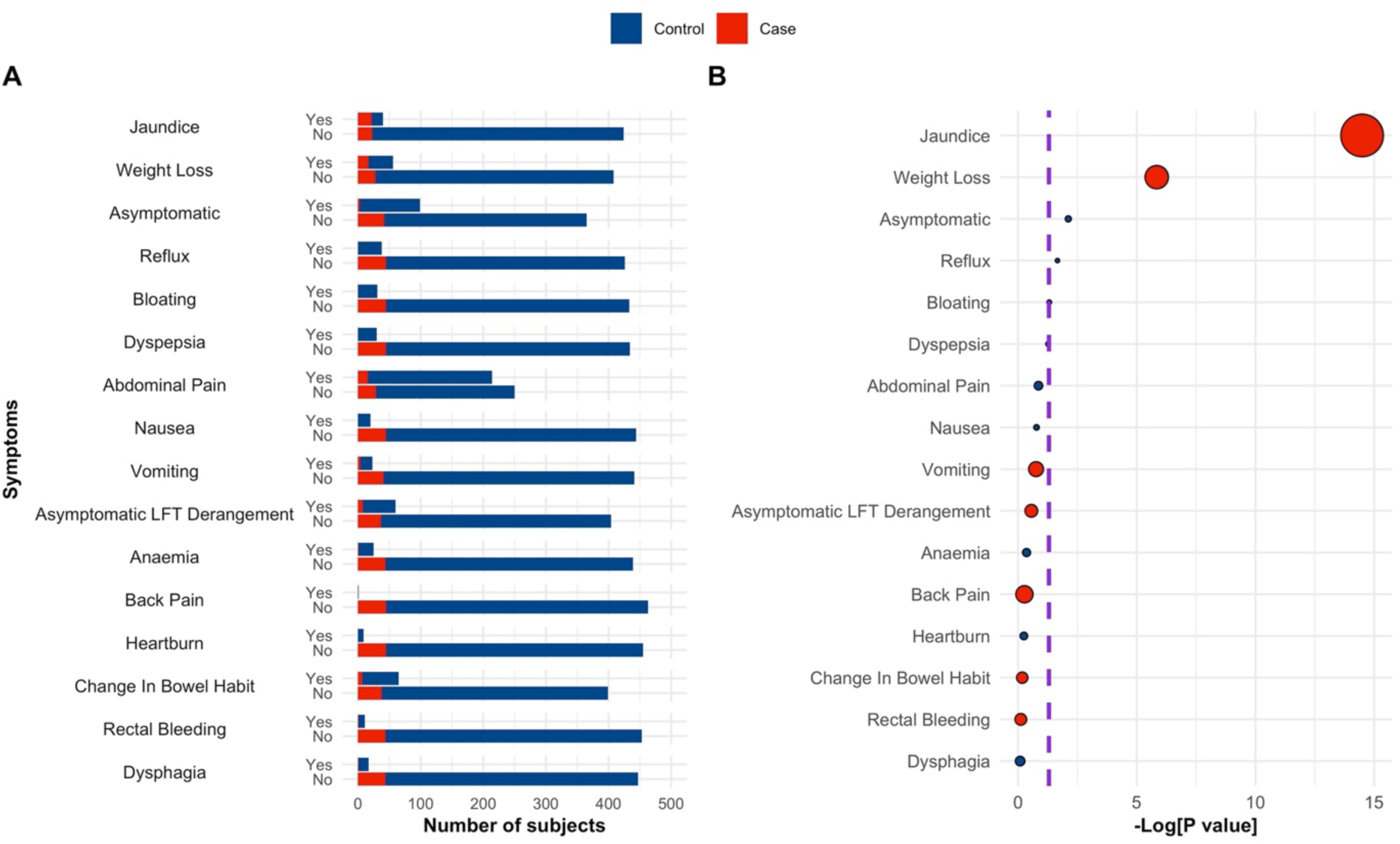
Association between symptoms and PDAC. **A** Number of subjects with each symptom according to PDAC status, case or control**. B** Association of symptoms with PDAC status, p values were calculated according to a logistic regression model with a bias reduction method. Purple dashed lines correspond to -Log [0.05]. In B dot sizes correspond to odds ratios and are colour coded according to their respective values, i.e., blue if OR<1 and red if OR>1. See also Supplementary Table 9. Only samples belonging to the ADEPS cohort were used as no information about symptoms was available for the UKCTOCS set of samples.

Within the framework presented in preceding sections, our ensemble of classifiers was developed independently of symptomatic data. To assess the overall efficacy of our signature and its predictive capacity for PDAC, we scrutinized its performance on a subset of ADEPTS patients, belonging to both discovery and validation cohorts, manifesting with ‘Weight Loss’ (n=56) and ‘Jaundice’ (n=40) (refer to Figure 4 and Supplementary Table 9). For each sample within these cohorts, where symptom data was accessible, probability scores were derived based on the ensemble model formulated using the whole discovery set presented above. We should emphasize that no additional model refinement was pursued. The decision to aggregate these probability scores is further rationalized by the relatively limited patient count exhibiting ‘Weight Loss’ and ‘Jaundice’ within the individual discovery and validation datasets (Supplementary Table 9). In the ADEPTS subset of samples presenting with ‘Weight Loss’, an AUC of 0.95 (95% CI 0.90 - 0.1), a sensitivity of 0.94 (95% CI 0.29 - 1), a PPV of 0.80 (95% CI 0.56 - 0.81), and a NPV of 0.97 (95% CI 0.74 - 1) at 90% specificity were achieved (Table 2 and Figure 5). In patients presenting with ‘Jaundice’, an AUC of 0.89 (95% CI 0.79 - 0.99), a sensitivity of 0.73 (95% CI 0.36 - 0.91), a PPV of 0.90 (95% CI 0.82 - 0.92), and a NPV of 0.73 (95% CI 0.54 - 0.89), at 90% specificity, were observed (Table 2 and Figure 5). Compared with the AUC obtained with a simple CA19-9 logistic regression model developed in the discovery set and by concatenating the probability scores in the discovery and validation as done above, a significantly lower AUC of 0.74 (95% CI 0.58 - 0.90) is achieved (p=1.29×10^-11^, one-sided bootstrap test), with a sensitivity of 0.53 (95% CI 0.24 - 0.76), a PPV of 0.70 (95% CI 0.50 - 0.77), and a NPV of 0.81 (95% CI 0.73 - 0.90), at 90% specificity, for patients presenting with ‘Weight Loss’. For patients presenting with ‘Jaundice’ an AUC of 0.70 (95% CI 0.53 - 0.86) is reached, also significantly inferior (p= 1.94×10^-7^), with a sensitivity of 0.41 (95% CI 0.14 - 0.73), a PPV of 0.83 (95% CI 0.64 - 0.90), and a NPV of 0.55 (95% CI 0.46 - 0.73), at 90% specificity, for CA19-9 as the single predictor.

**Table 2.** Performance summary for selected models in symptomatic patients. The probability values used to calculate the performance metrics were generated with each model developed in the training set and reported in the main text. Probability values for symptomatic patients belonging to the training set and validation set were concatenated to generate the ROC curves. Only ADEPTS samples had symptoms information. A. L. Derang.: Asymptomatic LFT Derangement. B. Pain: Back Pain. C. B. Habit: Change in Bowel Habit. W. Loss: Weight Loss. 95% confidence intervals are provided in parentheses. See also Supplementary Table 10 for the explicit performance ranks according to model, symptom and metric and Figure 5.

| Models | Metric | Symptom (Yes) |  |  |  |  |
| --- | --- | --- | --- | --- | --- | --- |
|  |  | A.L.Derang. | A. Pain | A. B. Habit | W.Loss | Jaundice |
| CA19-9 | ROC | 0.85 (0.62-1) | 0.81 (0.65-0.96) | 0.82 (0.57-1) | 0.74 (0.58-0.90) | 0.70 (0.53-0.86) |
|  | Sens90 | 0.75 (0.38-1) | 0.69 (0.44-0.94) | 0.71 (0.42-1) | 0.53 (0.24-0.76) | 0.41 (0.14-0.73) |
|  | PPV90 | 0.54 (0.37-0.61) | 0.36 (0.26-0.43) | 0.46 (0.34-0.55) | 0.70 (0.50-0.77) | 0.83 (0.64-0.90) |
|  | NPV90 | 0.96 (0.90-1) | 0.97 (0.95-0.99) | 0.96 (0.93-1) | 0.81 (0.73-0.90) | 0.55 (0.46-0.73) |
| Index signature | ROC | 0.98 (0.95-1) | 0.98 (0.97-1) | 0.97 (0.92-1) | 0.95 (0.90-1) | 0.89 (0.79-0.99) |
|  | Sens90 | 1 (0.62-1) | 0.94 (0.81-1) | 0.86 (0.56-1) | 0.94 (0.29-1) | 0.73 (0.36-0.91) |
|  | PPV90 | 0.61 (0.49-0.61) | 0.43 (0.40-0.45) | 0.51 (0.41-0.55) | 0.8 (0.56-0.81) | 0.90 (0.82-0.92) |
|  | NPV90 | 1 (0.94-1) | 0.99 (0.98-1) | 0.98 (0.95-1) | 0.97 (0.75-1) | 0.73 (0.54-0.89) |
| Reduced signature | ROC | 0.97 (0.93-1) | 0.92 (0.88-0.99) | 0.91 (0.83-0.98) | 0.92 (0.85-0.99) | 0.82 (0.67-0.97) |
|  | Sens90 | 1 (0.63-1) | 0.81 (0.38-1) | 0.57 (0.14-1) | 0.71 (0.35-0.94) | 0.77 (0-0.95) |
|  | PPV90 | 0.61 (0.49-0.94) | 0.40 (0.23-0.45) | 0.41 (0.15-0.55) | 0.75 (0.6-0.8) | 0.90 (0-0.92) |
|  | NPV90 | 1(0.94-1) | 0.98 (0.95-1) | 0.95 (0.9-1) | 0.88 (0.76-0.97) | 0.76 (0.42-0.94) |

**Figure 5.**
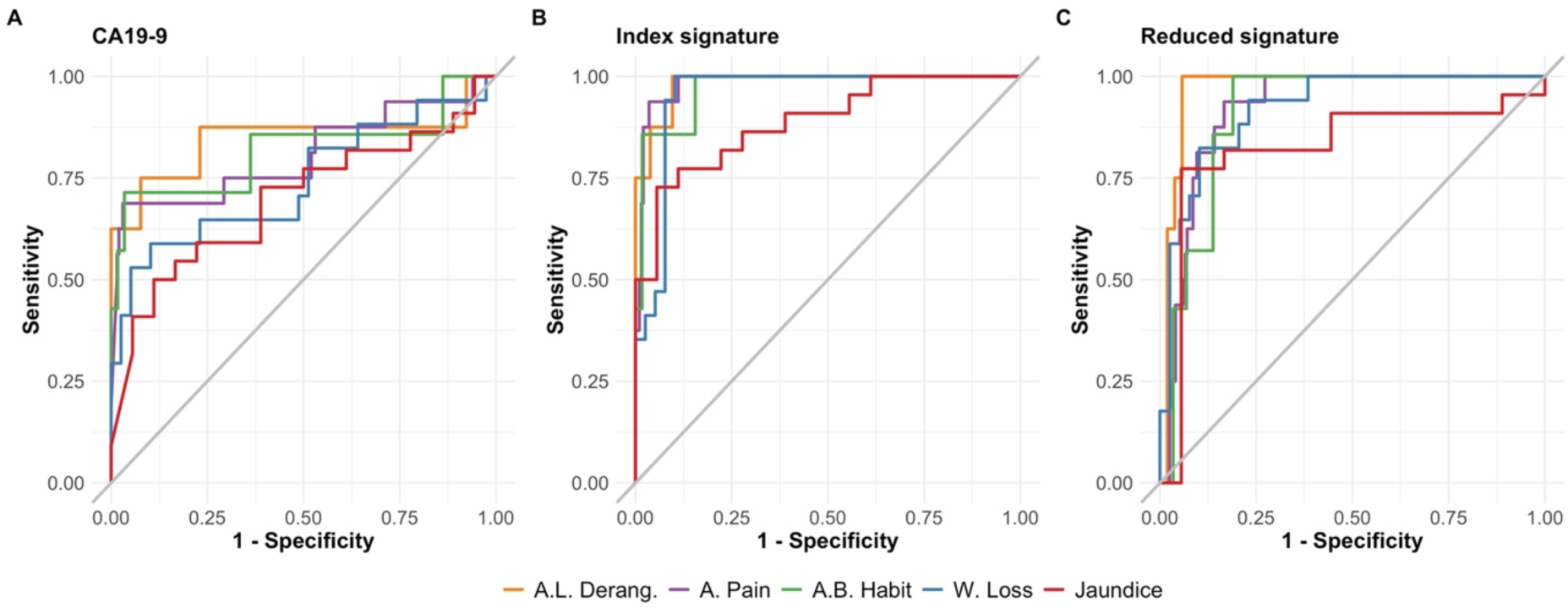
Receiver operating curves for selected models in symptomatic patients. **A** Only CA19-9. **B** Full index signature. **C** Reduced index signature. The probability values used to calculate the performance metrics were generated with each model developed in the discovery set and reported in the main text. Probability values for symptomatic patients belonging to the discovery set and validation set were concatenated to generate the ROC curves. Only ADEPTS samples had symptoms information. A. L. Derang.: Asymptomatic LFT Derangement. B. Pain: Back Pain. C. B. Habit: Change in Bowel Habit. W. Loss: Weight Loss. See also Table 2 for numerical values for area under the curve and other metrics.

With respect to other non-localising symptoms of note (Figure 5, Table 2 and Supplementary Table 10), the best predictive performance was noted for the full index signature where it was able to differentiate patients presenting with ‘abdominal pain’ due to benign conditions vs. PDAC with an AUC of 0.98 (95% CI 0.97-1), sensitivity of 0.94 (95% CI 0.81-1), PPV 0.43 (95% CI 0.40-0.45) and a NPV of 0.99 (95% CI 0.98-1), at 90% specificity. In those presenting with ‘change in bowel habit’, an AUC of 0.97 (95% CI 0.92-1), sensitivity 0.86 (95% CI 0.81-1), PPV of 0.51 (95% CI 0.41-0.55) and NPV of 0.98 (95% CI 0.95-1) was obtained. Both the index and the 8-marker signature showed superior predictive performance to CA19-9 as a single marker (see Supplementary Table 11 for the respective p-values).

### Correlation of the full PDAC ensemble signature with QCancer pancreatic score

In our final analysis, we juxtaposed the performance of our full ensemble classifier PDAC index against the QCancer risk prediction index, a clinical decision support tool available for primary care physicians, that integrates a myriad of individual-specific risk factors including age, sex, ethnicity, clinical measurements, diagnoses, and patient-reported symptoms into a risk stratifying point of care questionnaire (47). The ‘Today’s QCancer’ index evaluates an individual’s current risk of having an undiagnosed cancer as well as the specific risk for 9 distinct underlying cancer types, including pancreatic (‘pancreatic’ score) (56, 57). The aim was to determine whether in combination, the QCancer eCDST and our biomarker index signature would be able to better discriminate PDAC patients in a symptomatic (ADEPTS) cohort or whether it would be redundant. As the current risk threshold set by the NICE is at 3% for triggering specialist referrals (24), we opted to assess the combined performance of our index signature and the eCDST at a same or lower cut-off values.

The number of samples for which a QCancer score was computed is illustrated in Figure 6C. Using the diagnostic-specific ensemble model delineated previously, probability scores for samples in both discovery and validation cohorts were used to ascertain the combined ROC AUC for those samples possessing a QCancer score. This amalgamation was imperative, considering the reduced number of samples with an associated QCancer score (Figure 6C). It should be emphasized that no subsequent refinements or training of the algorithm were conducted. The ensemble stack index demonstrated a remarkable performance, achieving an AUC of 0.98 (95% CI 0.97 – 0.99), a sensitivity of 0.99 (95% CI 0.97-1), a PPV of 0.91 (95% CI 0.90 – 0.91), and a NPV of 0.99 (95% CI 0.96 - 1), at 90% specificity. Interestingly, when considering only samples with a QCancer risk above 2 or 2.5, the biomarker and clinical covariate ensemble index exhibited comparatively lower performance (Figure 6A). For a QCancer risk above 3.0, the performance of the index decreases minimally once again, which is expected as the difficulty of correctly singling out cases from confounding controls is increased (Figure 6A). However, the QCancer pancreatic score did exhibit a correlation with the odds of PDAC as determined by the ensemble classifier (R=0.36, p=3.4×10^-8^, Figure 6D) which highlights an important link between the purely clinical variables recorded for this cohort and the PDAC signature. Most importantly, the stacked index succeeded in attributing higher odds ratios above 1 to several PDAC cases that would have otherwise escaped detection had a QCancer score above 3 been taken as the risk predictor (Figure 6D). Contrarily, when depending exclusively on the QCancer score, and using it to calculate the ROC AUC, the predictive capacity for PDAC in the ADEPTS samples is noticeably diminished in comparison to the performance of the ensemble index (Figure 6B); this was verified in all samples with a Qcancer score (p=1.56×10^-18^, one-sided bootstrap test comparing AUCs), with a score above 2 (p=1.24×10^-10^), above 2.5 (p=3.68×10^-13^) and above 3 (p=2.33×10^-8^). This justifies the PDAC signature as a useful complementary resource for enhanced and accelerated diagnosis in the clinic.

**Figure 6.**
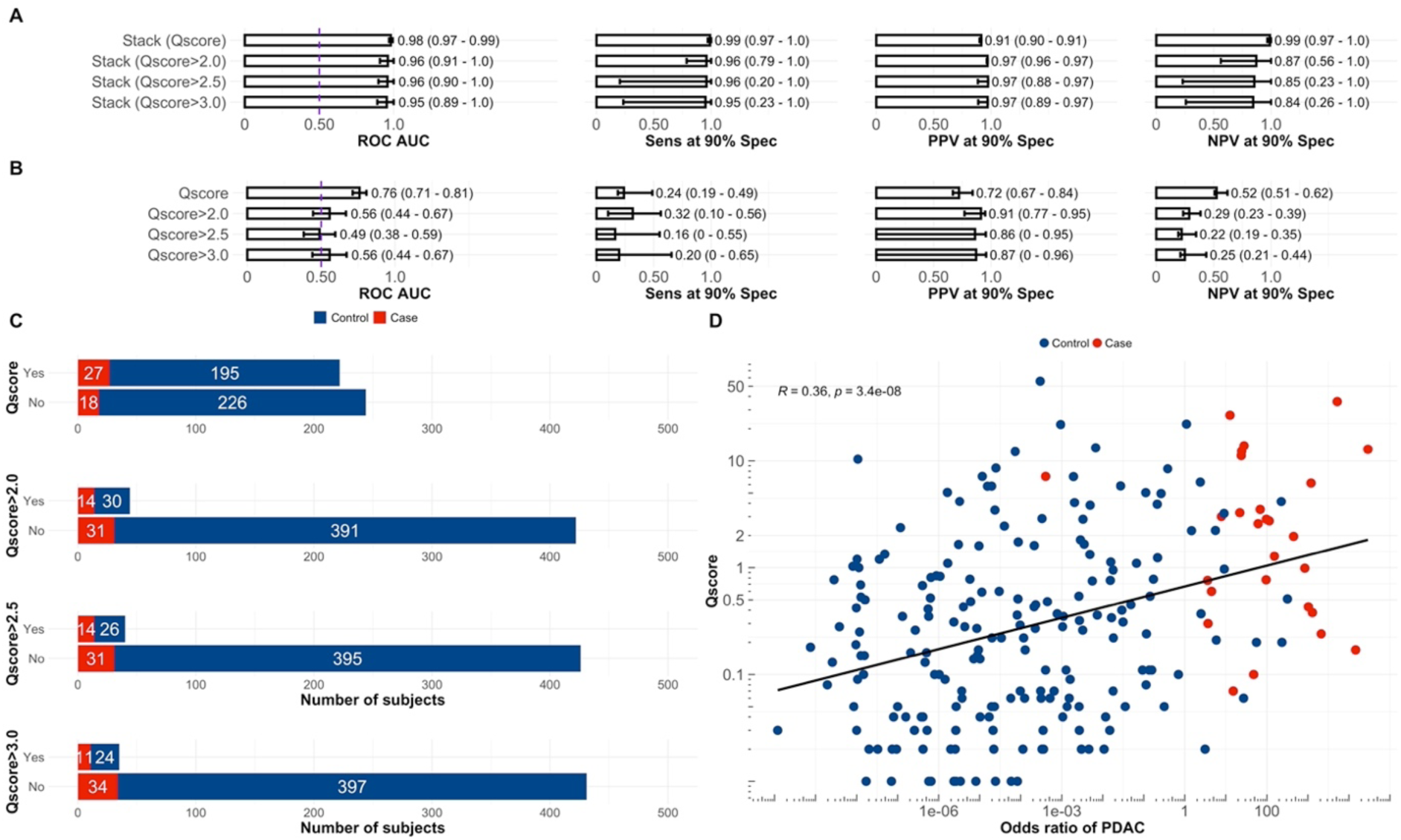
Prediction of PDAC in patients with specific symptoms and according to QCancer score values. The ensemble stack was selected as the best model according to Figure 2. **A** Performance of the stack in participants for which a Qscore had been calculated or above a specific threshold, bigger than 2, 2.5 or 3.0. **B** Performance of the Qscore taken as the predictor of PDAC risk in participants for which a Qscore had been calculated or above a specific threshold, bigger than 2, 2.5 or 3.0. **C** Number of subjects that had a calculated Qscore or are above a specific threshold, bigger than 2, 2.5 or 3.0**. D** Correlation between QCancer score and odds ratio of PDAC according to the stacked ensemble. E is in log scale. The QCancer score is identified as Qscore in the figure panels.

## Discussion

Our objective was to construct a multi-biomarker signature that could effectively differentiate individuals with non-specific yet concerning symptoms attributable to both benign abdominal pathologies and PDAC. CA19-9 tumour marker blood levels are used clinically to help confirm PDAC diagnosis in a clinical context (positive findings on imaging, histopathology), prognosticate and monitor recurrence following tumour resections (58). Its absent expression in Lewis body negative blood group individuals, an overall limited predictive capacity (79–81% test sensitivity and 82–90% specificity at best) and especially in the presence of certain inflammatory pancreatico-biliary conditions, have driven researchers to rather combine it in multi-marker panels to enhance its predictive performance (29, 30, 58). In an evolving multi-omics area, reported panels have included proteins, circulating nucleic acids (micro-RNA, cfDNA) or tumours cells, metabolites, and products of alternative DNA splicing and methylations (58, 59), developed to differentiate PDAC from healthy and those with benign pathologies. Yet, the role of such diagnostic and screening panels in symptomatic cohorts remains unestablished.

The sampled population in our study is an enriched, symptomatic, secondary care cohort where the prevalence of PDAC was close to 8%, representing figures observed in our hepatobiliary specialised referral centres. By using this target population and their unique set of serum samples provided by the ADEPTS study (33), we were able to develop a biomarker signature in a cohort of patients who were referred to our participating centres (University College London Hospitals, London UK and the Royal Free Hospital, London UK) with various abdominal and hepatobiliary conditions which in symptomatic presentation might overlap with PDAC (20). Moreover, we included samples from patients with known risk factors for PC (chronic pancreatitis, those with family history of PDAC and cystic lesions of the pancreas, CLPs) and with biliary conditions that are known confounders of CA19-9 (i.e. biliary tract inflammation/obstruction, pancreatitis, CLPs) - the only tumour marker clinically applied in the workup and management (29, 39, 60) of PDAC.

We employed ensemble methods, which have achieved impressive accuracy in numerous complex classification tasks (36, 42, 43, 61). Specifically, we utilized stacking—a form of meta-learning (43)—to create a superior-level predictive model based on the predictions of diagnostic-specific base classifiers. These classifiers leveraged a diverse set of features, highlighting the fundamental importance of heterogeneity arising from specific diagnoses when compared against PDAC, an approach previously demonstrated to be effective (36, 42). Moreover, this study enabled us to evaluate the specificity of our early detection machine learning approach (36) within a relevant symptomatic population, thereby allowing us to address confounding factors that may impact their performance. The use of such diagnostic specialized base-learners was further justified by the data asymmetry between PDAC cases and controls observed in both the discovery and validation datasets.

Across all diagnosis classes (base learners) the index signature which comprised 44 clinical and serum protein covariates predicted PDAC (all stages) with an AUC of 0.98 (95% CI 0.98 – 0.99); at 90% specificity, a sensitivity of 0.99 (95% CI 0.98 - 1), PPV 0.92 (95% CI 0.91 - 0.92) and NPV 0.99 (95% CI 0.97 - 1) was reached, in contrast to CA19-9 as a single predictor under a logistic regression model - AUC 0.79 (95% CI 0.66 – 0.91), sensitivity 0.67 (95% CI 0.50 - 0.83), PPV 0.32 (95% CI 0.26 - 0.38) and NPV of 0.97 (95% CI 0.96 - 0.99). On validation, an AUC of 0.95 (95% CI 0.91 – 0.99), sensitivity 0.86 (95% CI 0.68 - 1), PPV 0.54 (95% CI 0.48 - 0.58) and NPV of 0.98 (95% CI 0.95 - 1) was achieved by the signature, compared to an AUC of 0.80 (95% CI 0.66 – 0.93), sensitivity 0.65 (95% CI 0.48 - 0.56), PPV 0.49 (95% CI 0.41 - 0.56) and NPV of 0.95 (95% CI 0.92 - 0.98), for CA19-9.

The performance of this index panel must be appreciated within the context of the complex biology associated with each of the ensembled diagnostic classes, i.e., the challenges associated with biomarker alterations on the background of pancreatico-biliary inflammatory and obstructive pathologies. When applying a redacted, 8-marker signature (CA19-9, VWF, CPE, CTSV, CEACAM1,CD160, Diabetes and Age) - features that were selected with relatively high importance across most base learners, the performance was naturally reduced, yet still performed significantly better against CA19-9 as a single marker during discovery. Using the general linear model stack as was done in the case of the full index, the reduced signature predicted PDAC with AUC of 0.97 (95% CI 0.95-0.98), sensitivity 0.98 (95% CI 0.95-1), PPV 0.92 (95% CI 0.91-0.92) and NPV of 0.98 (95% CI 0.94-1), at 90% specificity (Supplementary Figure 4C), values comparable to the full index. During validation, however, the predictive capacity of the reduced signature was significantly reduced compared to the full stacked model (Supplementary Figure 4D) and only marginally superior to CA19-9 alone across the cohort. In contrast, it still outperformed CA19-9 by a significant margin when predicting PDAC against healthy controls.

As validation of its performance, we also applied the full index signature to the cohort, which were re-stratified based on presenting symptoms in contrast to the ensemble of classifiers which were developed independently of symptomatic data and with no further model refinement. The aim was to test the signature performance in differentiating PDAC cases from controls by accounting for presenting symptoms, and which have been linked with repeated primary care consultations up to two-years prior to PDAC diagnosis (20) (Figure 4). Enriched by fulfilling certain sociodemographic, clinical and attributable suspicious symptom (identified using CDSTs such as QCancer tool), symptomatic patients would form an ideal cohort for further risk stratification by minimally invasive blood biomarker testing for prioritisation of more invasive (and costly) investigations. Yet, contrary to the full index, in the cohort used in the current work the QCancer score used as the sole predictor of PDAC did not achieve significant performances in samples above the threshold of 3%. This further motivates the recourse to combined strategies where complementary biomarker panels such as those identified by ensemble modelling approaches could improve early detection when used in conjunction with CDSTs.

In our test subjects, however, only ‘Jaundice’ (p=3.22×10^-15^), and ‘Weight Loss’ (p=1.44×10^-6^) were significantly associated with PDAC. When testing the diagnostic performance of the full index signature in all symptomatic patients presenting with ‘Weight Loss’, the signature significantly outperformed CA19-9: AUC_signature_ of 0.95 (95% CI 0.90 - 0.1) vs. AUC_CA19-9_ of 0.74 (95% CI 0.58 - 0.90) (Figure 5A, Table 2 and Supplementary Table 11). ‘Weight loss’ has previously been reported to have the longest diagnostic interval in a prospective primary cohort study (SYMPTOM pancreatic study), assessing symptom trends and associated diagnostic intervals in PC (14). Attesting to the full index signature’s capacity as a rule out test in such patients, is its outstanding negative predictive value compared to that of CA19-9 (0.97 95% CI 0.75-1 vs. 0.81 95% CI 0.73-0.9, respectively) (Table 2). Similarly, the index signature performed superiorly to CA19-9 in jaundiced patients (AUC of 0.89 (95% CI 0.79 - 0.99) vs. CA19-9 AUC of 0.70 (95% CI 0.53 - 0.86), see also Supplementary Table 11), which underscores once again the increased capacity of the ensemble index to better identify PDAC in the presence of a known confounder of CA19-9 (29, 39).

While our study provides valuable insights, it is not without limitations. While the observed prevalence of PDAC in this study aligns with secondary care population trends, enhanced specificity and positive predictive value would necessitate larger cohorts with an increased number of cases. Moreover, the sample set representing the ‘Healthy’ control class warrants expansion to incorporate a more diverse population of both men and women. This control class, derived from the UKCTOCS samples used in a previous study (36), exclusively comprised women. Given its superior performance in predicting PDAC, as depicted in Figure 2, the inclusion of male samples within this class could further enhance the breadth of the panel of markers identified in this study. Lastly, although diabetes emerged both as a risk factor and a central clinical covariate in our signature (including in the reduced panel), we must emphasize and recognise the lack of complete (type, duration) data in the UKCTOCS cohort (36). Nevertheless, diabetes mellitus (and in particularly of new onset) is an established risk factor and therefore its inclusion as a relevant feature in the signature is of no surprise (9).

While both our index and reduced signature were superior to CA19-9 in their predictive performance and compensated for asymmetric binary classes by creating a diagnostic-specific ensemble, its complexity challenges its utilisation in clinic. Yet, in the current era of rapidly evolving assay technologies, the utilization of a complex biomarker signature comprising numerous variables has gained significant relevance. While the complexity of these biomarker signatures may pose analytical challenges, the evolving assay technologies offer the means to effectively harness their potential.

Future enhancements however, will naturally necessitate the study of larger cohorts, potentially incorporating a biomarker-contextualized machine learning perspective that accounts for sample-specific aspects related to diagnosis, a strategy employed in other cancer research domains (62). The utilization of disease trajectory tracking and clinical history analysis (63) may also facilitate the application of advanced deep learning techniques and electronic health data. When combined with ensemble biomarker signatures taken for example in a longitudinal context (36, 64), these approaches could enhance the estimation of PDAC risk within an enriched symptomatic population.

## Data Availability

Data requestors will need to sign a data access agreement and in keeping with patient consent for secondary use obtain ethical approval for any new analyses.

## Acknowledgments

We thank the participants of the ADEPTS study and UKCTOCS trial, the management team, research nurses, interviewers, research assistants, and other staff who gathered the data that was used in this work. This research was funded by Cancer Research UK (grant C12077/A26223) and supported by the Pancreatic Cancer UK Early Diagnosis Award 2018, project “The Accelerated Diagnosis of neuroEndocrine and Pancreatic TumourS (ADEPTS)” (IRAS Number: 234637, NIHR Portfolio no. 7343), and by the National Institute for Health Research (NIHR) University College London Hospitals Biomedical Research Centre. UKCTOCS was core funded by the Medical Research Council, Cancer Research UK, and the Department of Health with additional support from the Eve Appeal, Special Trustees of Bart’s and the London, and Special Trustees of University College London Hospitals. EC is supported by Cancer Research UK (C7690/A26881). UM acknowledges MRC Core funding (MC_UU_00004/01). SPP is supported by Cancer Research UK Early Detection and Diagnosis Programme EDDPGM-May22\100002 (CANDETECT, Co-PI Prof Fiona Walter).

## Author contributions

AN, NRN, SPP, UM and AZ conceived the study. SPP, AZ and UM secured funding. NRN constructed the models, performed the statistical analysis and produced the figures. AN and NRN interpreted the data in collaboration with SPP and AZ. AN, NRN, SPP and AZ drafted the paper. ES and AN performed the experiments for all in-house biomarkers. All authors contributed to data acquisition and interpretation, and critically reviewed and approved the article: AN, NRN, ES, PA, OB, HJW, EC, AGM, NRW, UM, GKF, AZ, SPP.

## Supplementary Information

### Supplementary Figures

**Supplementary Figure 1.**
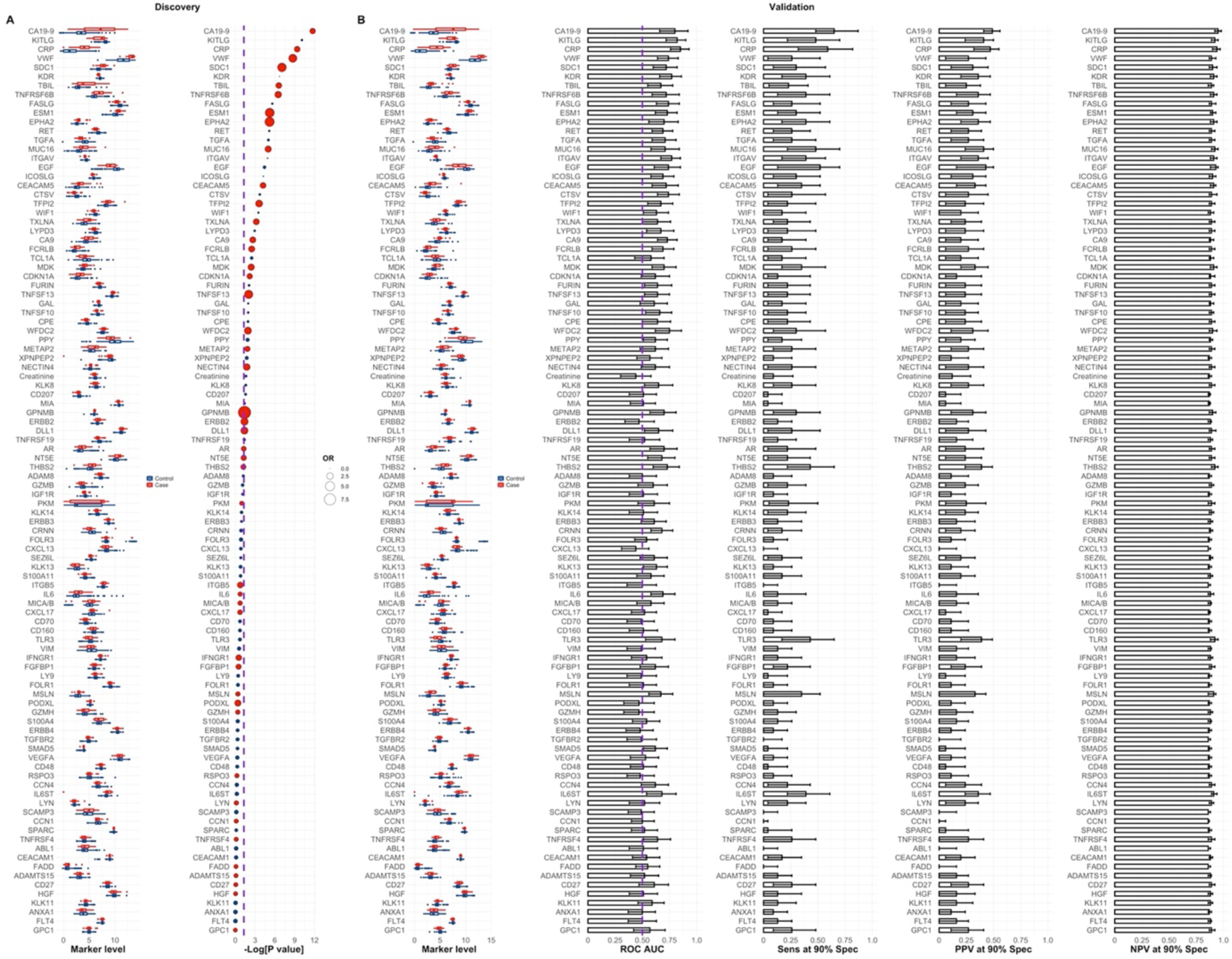
Biomarker ranks in the discovery set. **A** Distribution and ranks of biomarkers by p values in the discovery set. Purple dashed line corresponds to -Log [0.05]. **B** Receiver Operating Curve (ROC) Area Under the Curve (AUC), Sensitivity (Sens), Positive Predictive Value (PPV) and Negative Predictive Value (NPV) at 90% Specificity (Spec) performance of single marker models in the validation set set. OR stands for odds-ratio, with dot size proportional to the calculated values. Red and blue OR points represent OR > 1 (favours pancreatic ductal adenocarcinoma (PDAC) case status) and OR < 1 (favours Control status), respectively. p values were calculated according to a logistic regression model with a bias reduction method. Performances were calculated with the single feature models developed in the discovery set. The ROC AUC significance threshold is also represented by a purple dashed line at 0.5. Error bars in figures corresponding to the validation set correspond to 95% Confidence Intervals (CI), calculated by stratified bootstrapping 2000 times. See statistical analysis section in Methods (main text) for further details and Supplementary Table 3 and 4.

**Supplementary Figure 2.**
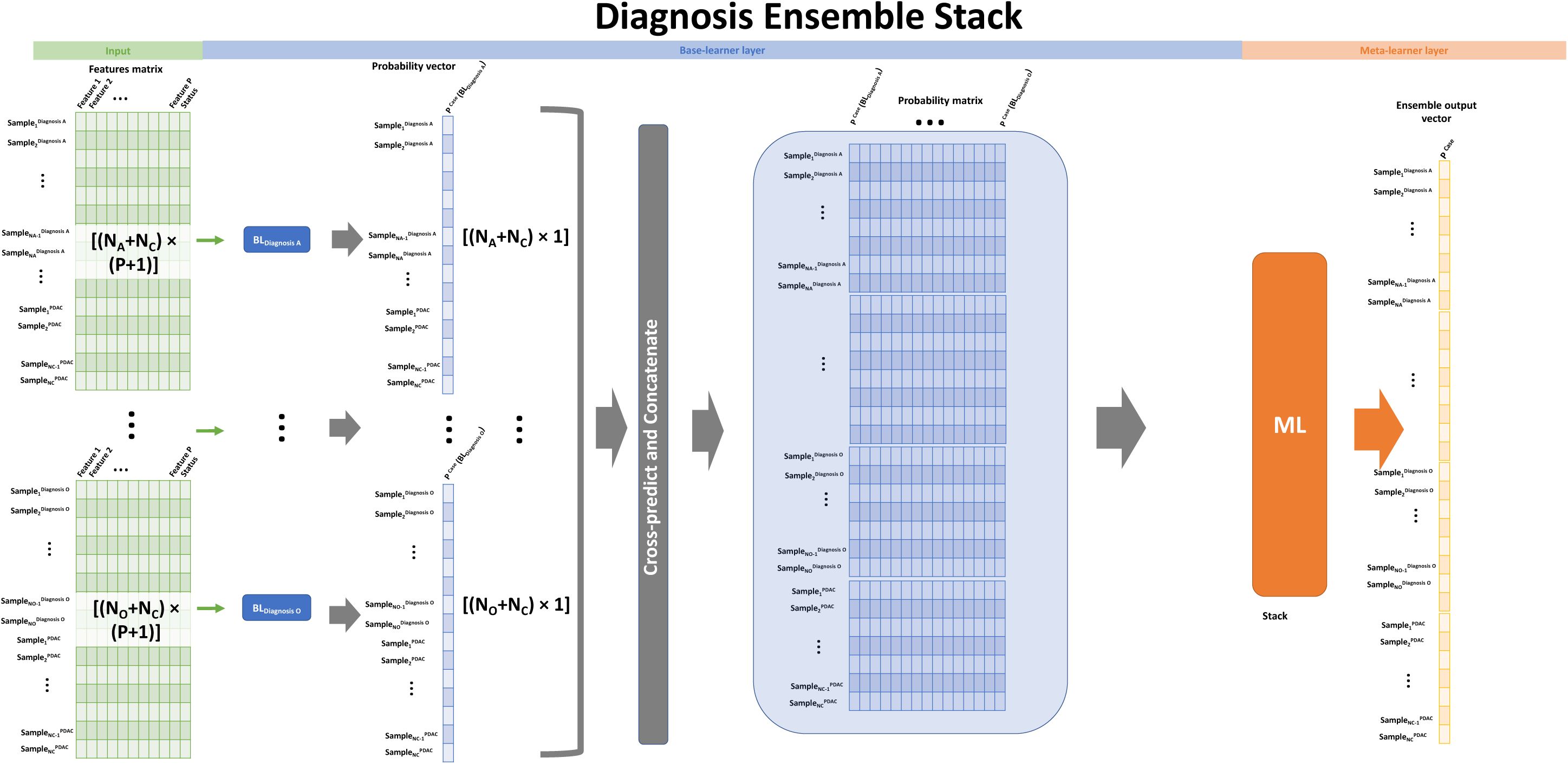
Flow diagram for the stack ensemble classifier. The general linear stack model presented in the main text was built according to this diagram, where base-learners are trained in groups of control samples with a specific diagnosis and the same PDAC cases. The stacking procedure has 2 steps. First, the probability output vectors for each base-learner are concatenated, thus leading to n probability vectors, where n is the number of diagnosis subclasses. Second, these vectors are subsequently used to populate the diagonal blocks of a large probability matrix. The off-diagonal probability blocks are generated by using the base models trained in a specific diagnosis subclass plus the same PDACs, which therefore amounts to computing cross diagnosis predictions. The resulting large matrix has n columns and is then used to train the meta-learner which outputs the final probability vector. For the purposes of applying the resulting trained models to the validation set, the flow of the diagram is the same as before, but the feature matrix will have a different number of samples.

**Supplementary Figure 3.**
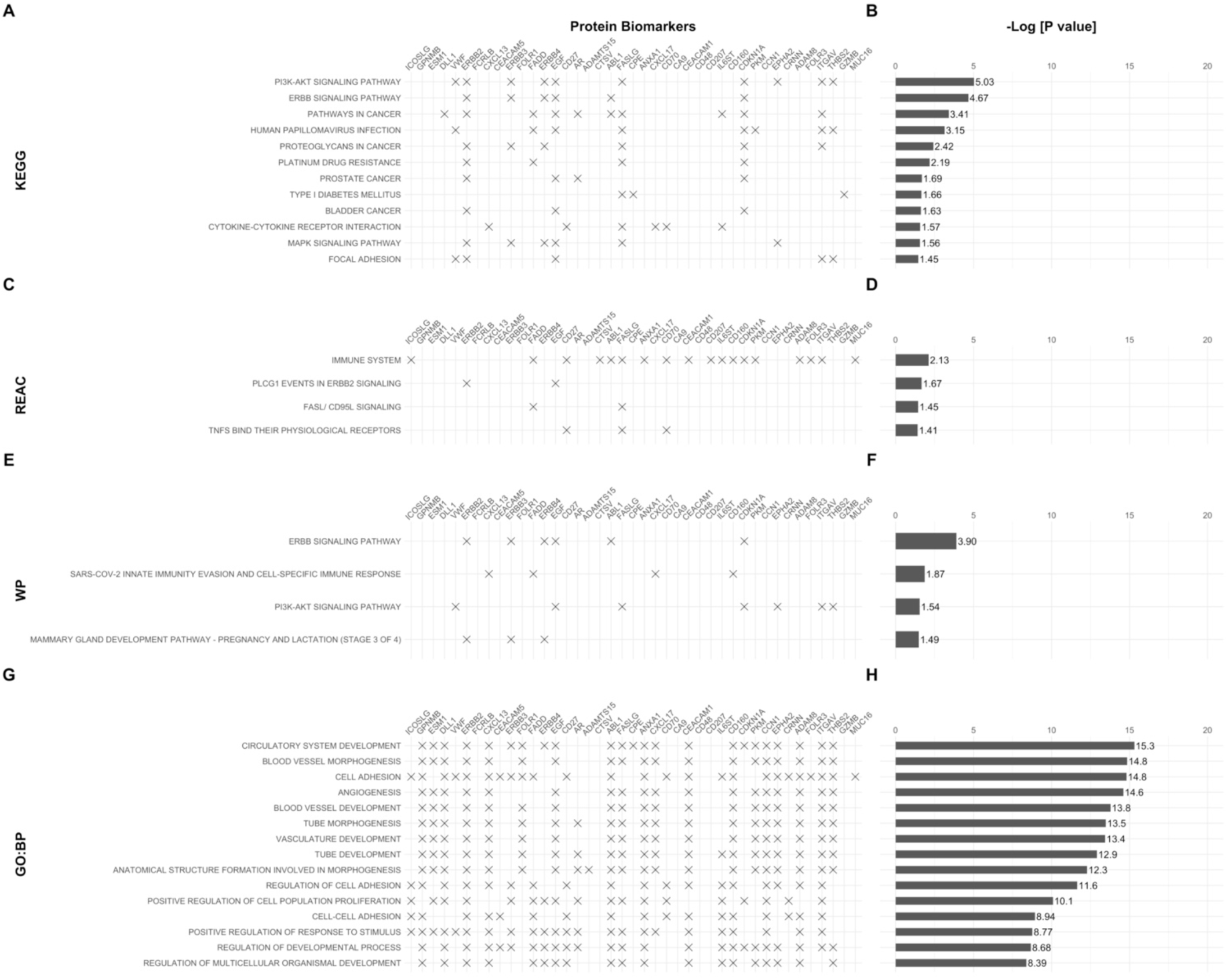
Enrichment analysis. g:Profiler terms for the set of features selected by the full stack ensemble. **A** Kyoto Encyclopaedia of Genes and Genomes (KEGG) pathways. **C** Reactome Pathway Database (REAC). **E** WikiPathways (WP). **G** Gene ontology terms biological process (GO: BP). The respective adjusted p-values associated with each enrichment term or pathway are plotted in **B**, **D**, **F** and **H**. See also Figure 3. See Statistical Analysis in Methods for further details.

**Supplementary Figure 4.**
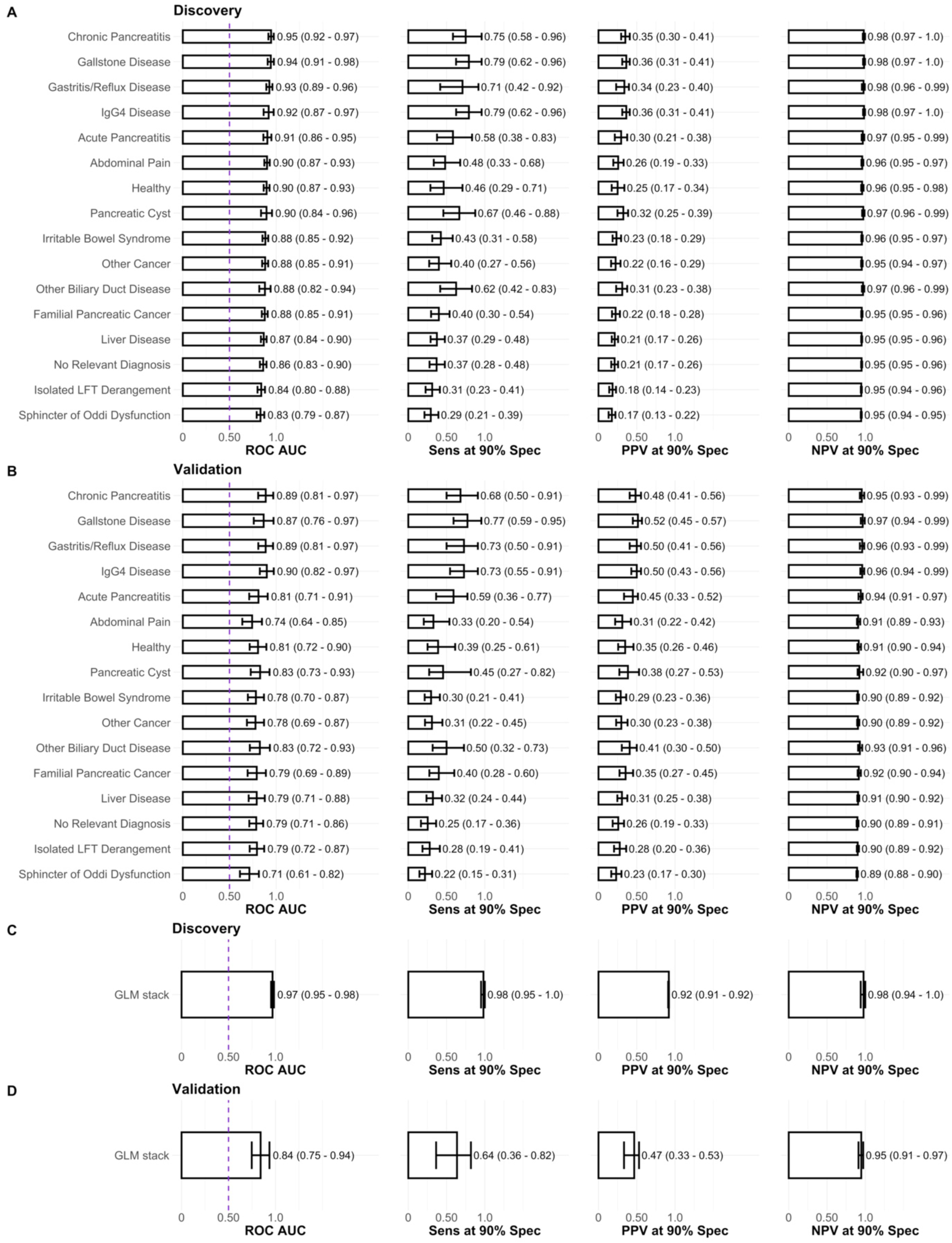
Performance of individual base-learner classifiers and stack ensemble for a reduced set of biomarkers, CA19-9, Age, Diabetes, VWF, CPE, CTSV, CEACAM1 and CD160. **A** Base-learner performance in the discovery. Each base-learner classifier was developed by training with a recursive feature elimination technique (RFE) and logistic regression (glm) in samples belonging to each specific diagnosis class against the same 24 PDACs in the discovery set. The performance reported in A is, nevertheless, of each classifier in the whole discovery set. The performances reported in **B** correspond to the base-learners developed in the discovery set but applied to the whole validation set. In **C** and **D** the performance of an ensemble GLM stack based on the base-learners presented in A and B is reported in the discovery and validation sets, respectively. The ROC AUC significance threshold is represented by a purple dashed line at 0.5. Error bars in figures correspond to 95% Confidence Intervals (CI), calculated by stratified bootstrapping 2000 times.

**Supplementary Figure 5.**
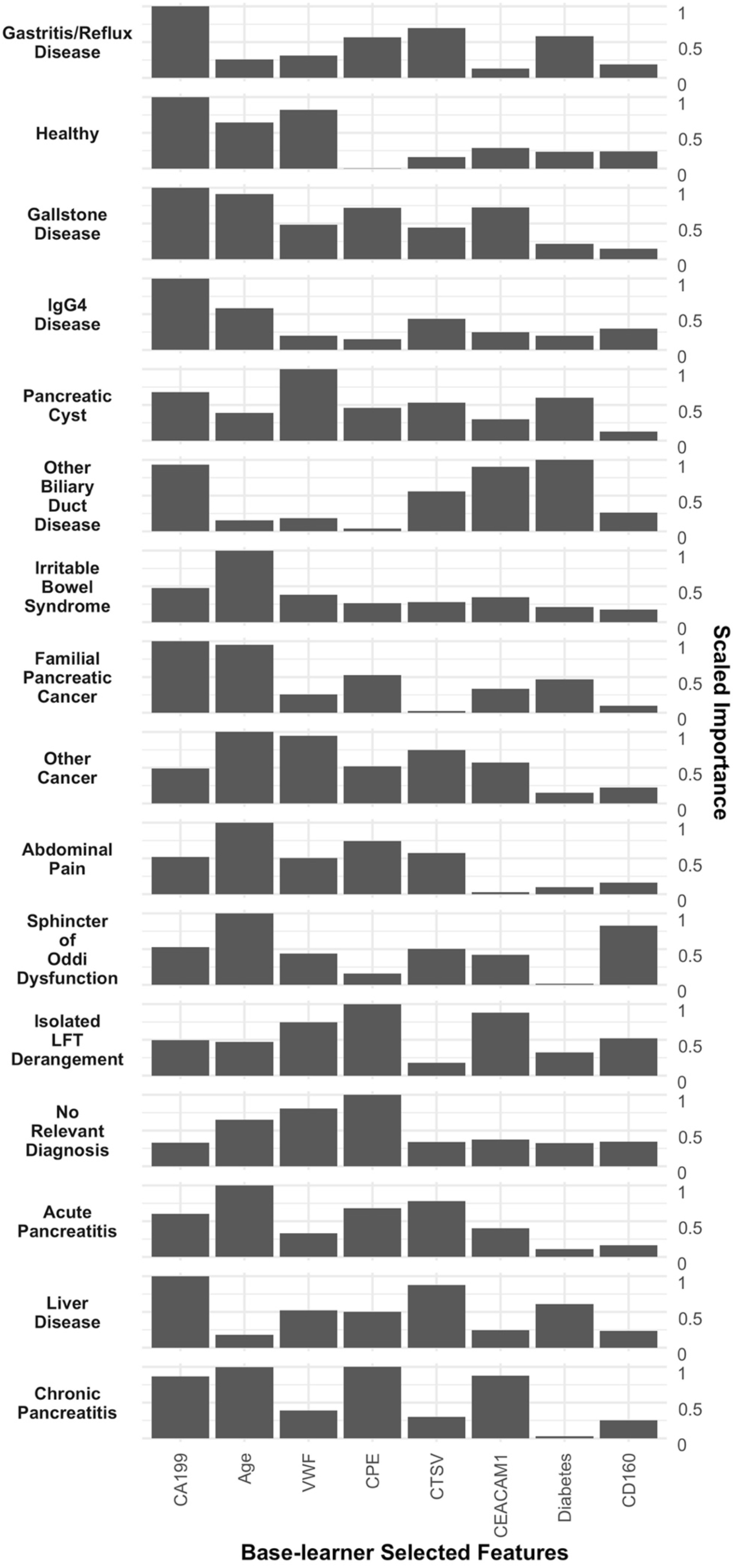
Feature importance for a reduced 8-marker model following the same principles as those described in Figure 2 A and B. The reduced set of biomarkers was CA19-9, Age, Diabetes, VWF, CPE, CTSV, CEACAM1 and CD160. See also performances in Supplementary Figure 4. Selected features are ranked from left to right according to the average scaled importance across base learners. See Methods for details.

**Supplementary Figure 6.**
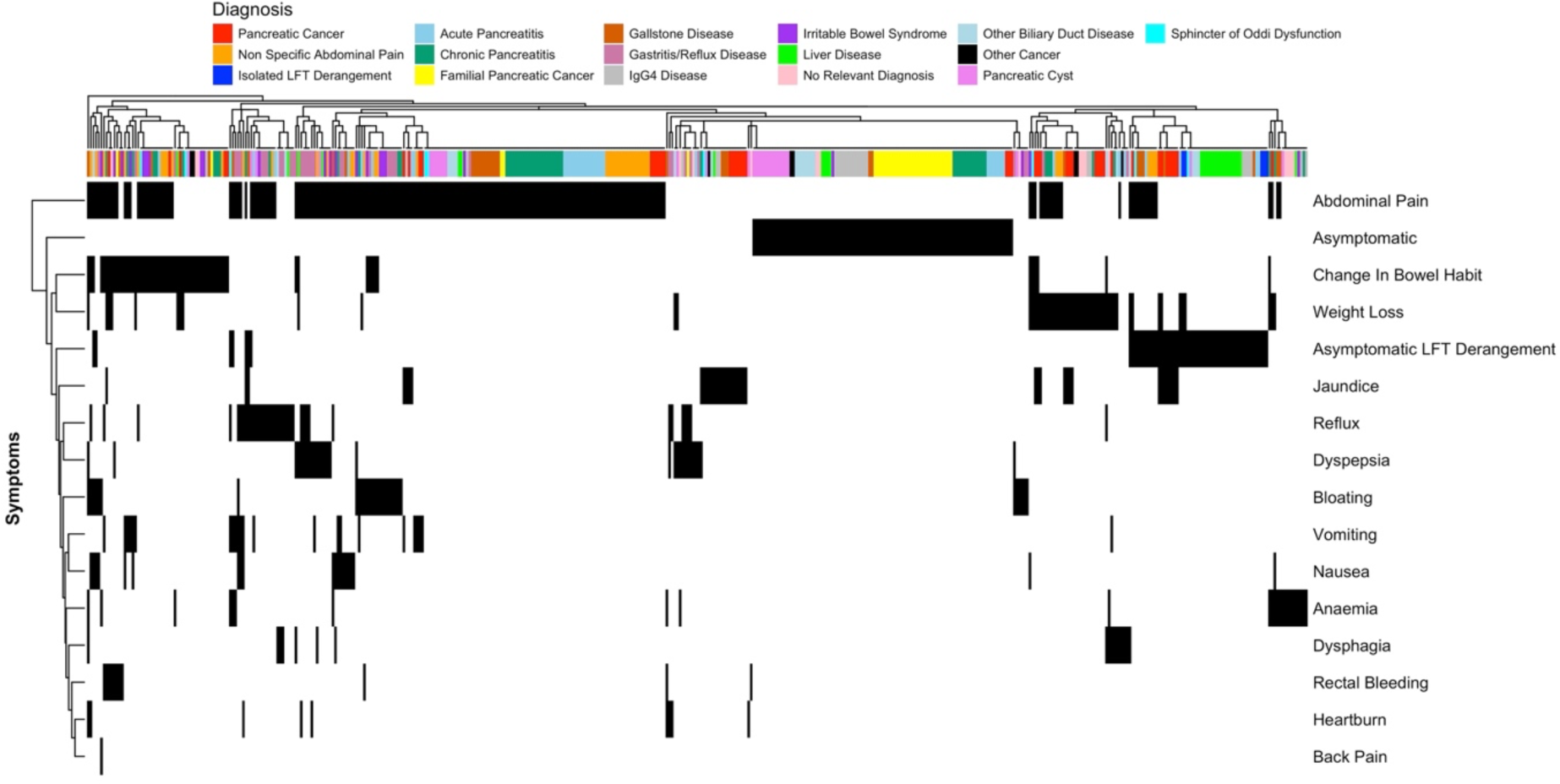
Matrix for associated symptoms for each sample. Each sample in the whole set collected from ADEPTS cohort is represented by the columns. The colour coding corresponds to the diagnosis class associated with each sample. Symptoms are represented in each row. Both columns and rows are clustered according to their symptoms pattern. Black represents presence of sample with the respective symptoms and diagnosis.

### Supplementary Tables

**Supplementary Table 1.** Quantitative ELISA assays’ intra-assay coefficient of variation.

| Assay | Dilution factor | CV(%) |
| --- | --- | --- |
| CA19-9(A) | 1:4 | 6.9 |
| VWF | 1:100 | 13.5 |
| THBS2 | 1:10 | 12.5 |
| PKM/PKM2 | 1:10 | 10.3 |
| IL6ST/IL6RB | 1:100 | - |

**Supplementary Table 2.**
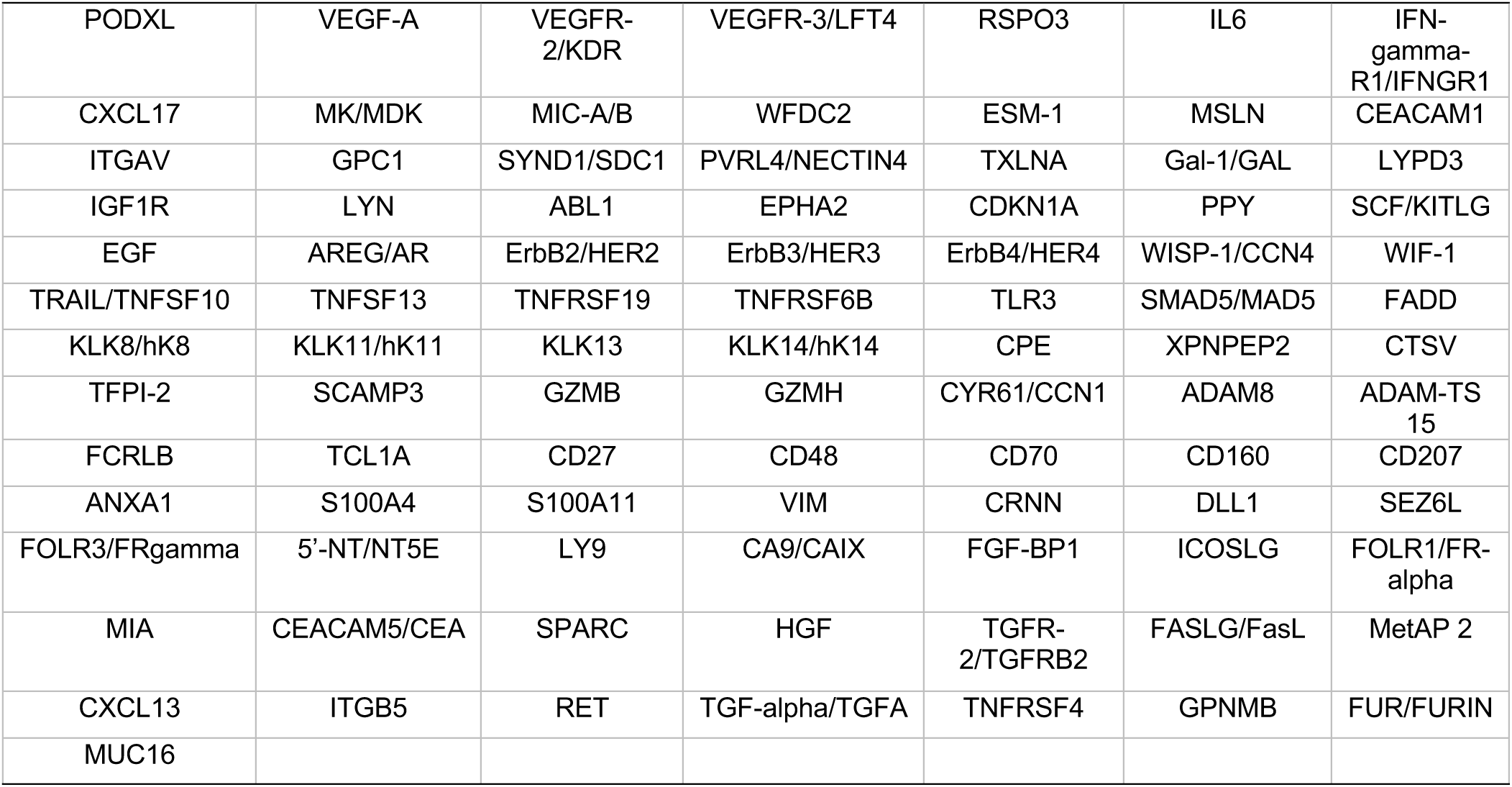
Cancer-associated proteins measured on the Olink Oncology II panel used in this project. The remaining biomarkers were done in-house. The protein names are listed to be consistent with those provided by Olink.

|  |  |  |  |  |  |  |
| --- | --- | --- | --- | --- | --- | --- |
| PODXL | VEGF-A | VEGFR-2/KDR | VEGFR-3/LFT4 | RSPO3 | IL6 | IFN-gamma-R1/IFNGR1 |
| CXCL17 | MK/MDK | MIC-A/B | WFDC2 | ESM-1 | MSLN | CEACAM1 |
| ITGAV | GPC1 | SYND1/SDC1 | PVRL4/NECTIN4 | TXLNA | Gal-1/GAL | LYPD3 |
| IGF1R | LYN | ABL1 | EPHA2 | CDKN1A | PPY | SCF/KITLG |
| EGF | AREG/AR | ErbB2/HER2 | ErbB3/HER3 | ErbB4/HER4 | WISP-1/CCN4 | WIF-1 |
| TRAIL/TNFSF10 | TNFSF13 | TNFRSF19 | TNFRSF6B | TLR3 | SMAD5/MAD5 | FADD |
| KLK8/hK8 | KLK11/hK11 | KLK13 | KLK14/hK14 | CPE | XPNPEP2 | CTSV |
| TFPI-2 | SCAMP3 | GZMB | GZMH | CYR61/CCN1 | ADAM8 | ADAM-TS 15 |
| FCRLB | TCL1A | CD27 | CD48 | CD70 | CD160 | CD207 |
| ANXA1 | S100A4 | S100A11 | VIM | CRNN | DLL1 | SEZ6L |
| FOLR3/FRgamma | 5'-NT/NT5E | LY9 | CA9/CAIX | FGF-BP1 | ICOSLG | FOLR1/FR-alpha |
| MIA | CEACAM5/CEA | SPARC | HGF | TGFR-2/TGFRB2 | FASLG/FasL | MetAP 2 |
| CXCL13 | ITGB5 | RET | TGF-alpha/TGFA | TNFRSF4 | GPNMB | FUR/FURIN |
| MUC16 |  |  |  |  |  |  |

**Supplementary Table 3.** Odds ratios, p values, ROC AUC, sensitivity (Sens), positive predictive value (PPV) and negative predictive value (NPV) for univariate logistic regression models derived in the discovery set. The performances of these model in the validation are presented in Supplementary Table 4. Markers are ranked according to the p values. Spec: specificity.

| Marker | OR | p value | ROC AUC | Sens at 90%<br>Spec | PPV at 90%<br>Spec | NPV at 90%<br>Spec |
| --- | --- | --- | --- | --- | --- | --- |
| CA19 - 9 | 1.85 (1.54 - 2.26) | 1.87E-12 | 0.79 (0.66 - 0.91) | 0.67 (0.5 - 0.83) | 0.32 (0.26 - 0.38) | 0.97 (0.96 - 0.99) |
| KITLG | 0.23 (0.14 - 0.37) | 7.95E-11 | 0.79 (0.68 - 0.89) | 0.5 (0.29 - 0.71) | 0.26 (0.17 - 0.34) | 0.96 (0.95 - 0.98) |
| CRP | 2.01 (1.59 - 2.61) | 4.12E-10 | 0.84 (0.74 - 0.92) | 0.58 (0.38 - 0.83) | 0.34 (0.25 - 0.42) | 0.96 (0.94 - 0.98) |
| VWF | 3.92 (2.28 - 7.78) | 1.89E-09 | 0.84 (0.76 - 0.9) | 0.5 (0.29 - 0.71) | 0.26 (0.17 - 0.34) | 0.96 (0.95 - 0.98) |
| SDC1 | 4.42 (2.56 - 8.01) | 8.47E-08 | 0.79 (0.66 - 0.89) | 0.67 (0.46 - 0.83) | 0.32 (0.25 - 0.38) | 0.97 (0.96 - 0.99) |
| KDR | 0.01 (0.00 - 0.07) | 2.02E-07 | 0.8 (0.72 - 0.87) | 0.42 (0.21 - 0.67) | 0.23 (0.13 - 0.32) | 0.96 (0.94 - 0.97) |
| TBIL | 2.03 (1.55 - 2.75) | 2.53E-07 | 0.72 (0.6 - 0.85) | 0.54 (0.29 - 0.71) | 0.32 (0.2 - 0.38) | 0.96 (0.94 - 0.97) |
| TNFRSF6B | 2.62 (1.80 - 3.94) | 3.12E-07 | 0.75 (0.64 - 0.86) | 0.5 (0.29 - 0.71) | 0.26 (0.17 - 0.34) | 0.96 (0.95 - 0.98) |
| FASLG | 0.20 (0.10 - 0.39) | 2.39E-06 | 0.77 (0.65 - 0.87) | 0.46 (0.21 - 0.67) | 0.25 (0.13 - 0.32) | 0.96 (0.94 - 0.97) |
| ESM-1 | 4.84 (2.35 - 10.69) | 5.99E-06 | 0.76 (0.66 - 0.84) | 0.29 (0.12 - 0.5) | 0.17 (0.08 - 0.27) | 0.95 (0.93 - 0.96) |
| EPHA2 | 5.15 (2.56 - 10.97) | 6.31E-06 | 0.73 (0.63 - 0.82) | 0.29 (0.12 - 0.46) | 0.17 (0.08 - 0.25) | 0.95 (0.93 - 0.96) |
| RET | 0.18 (0.08 - 0.38) | 7.01E-06 | 0.78 (0.68 - 0.87) | 0.42 (0.21 - 0.62) | 0.23 (0.13 - 0.31) | 0.96 (0.94 - 0.97) |
| TGFA | 0.33 (0.18 - 0.55) | 9.61E-06 | 0.79 (0.69 - 0.87) | 0.33 (0.17 - 0.62) | 0.19 (0.11 - 0.31) | 0.95 (0.94 - 0.97) |
| MUC16 | 2.39 (1.63 - 3.65) | 1.02E-05 | 0.72 (0.59 - 0.83) | 0.38 (0.21 - 0.58) | 0.21 (0.13 - 0.3) | 0.95 (0.94 - 0.97) |
| ITGAV | 0.05 (0.01 - 0.19) | 1.29E-05 | 0.78 (0.68 - 0.87) | 0.46 (0.17 - 0.67) | 0.25 (0.11 - 0.32) | 0.96 (0.94 - 0.97) |
| EGF | 0.55 (0.41 - 0.72) | 3.77E-05 | 0.79 (0.71 - 0.87) | 0.5 (0.21 - 0.71) | 0.26 (0.13 - 0.34) | 0.96 (0.94 - 0.98) |
| ICOSLG | 0.04 (0.01 - 0.20) | 5.60E-05 | 0.77 (0.66 - 0.85) | 0.29 (0.12 - 0.5) | 0.17 (0.08 - 0.26) | 0.95 (0.93 - 0.96) |
| CEACAM5 | 2.05 (1.46 - 2.94) | 6.15E-05 | 0.72 (0.61 - 0.83) | 0.33 (0.17 - 0.58) | 0.19 (0.11 - 0.3) | 0.95 (0.94 - 0.97) |
| CTSV | 0.27 (0.13 - 0.54) | 0.00019 | 0.73 (0.61 - 0.84) | 0.29 (0.08 - 0.5) | 0.17 (0.06 - 0.26) | 0.95 (0.93 - 0.96) |
| TFPI2 | 2.91 (1.65 - 5.38) | 0.00024 | 0.7 (0.58 - 0.8) | 0.29 (0.08 - 0.5) | 0.17 (0.06 - 0.26) | 0.95 (0.93 - 0.96) |
| WIF1 | 0.17 (0.06 - 0.45) | 0.00031 | 0.72 (0.61 - 0.82) | 0.33 (0.12 - 0.5) | 0.19 (0.08 - 0.26) | 0.95 (0.93 - 0.96) |
| TXLNA | 2.21 (1.41 - 3.51) | 0.00065 | 0.69 (0.55 - 0.81) | 0.42 (0.17 - 0.62) | 0.23 (0.11 - 0.31) | 0.96 (0.94 - 0.97) |
| LYPD3 | 0.24 (0.10 - 0.57) | 0.0012 | 0.68 (0.57 - 0.79) | 0.33 (0.17 - 0.58) | 0.19 (0.11 - 0.3) | 0.95 (0.94 - 0.97) |
| CA9 | 2.21 (1.33 - 3.68) | 0.0022 | 0.65 (0.52 - 0.77) | 0.29 (0.12 - 0.5) | 0.17 (0.08 - 0.26) | 0.95 (0.93 - 0.96) |
| FCRLB | 2.24 (1.32 - 3.74) | 0.0033 | 0.66 (0.53 - 0.78) | 0.38 (0.17 - 0.58) | 0.21 (0.11 - 0.3) | 0.95 (0.94 - 0.97) |
| TCL1A | 0.54 (0.35 - 0.82) | 0.0033 | 0.68 (0.57 - 0.78) | 0.29 (0.12 - 0.5) | 0.17 (0.08 - 0.26) | 0.95 (0.93 - 0.96) |
| MDK | 2.42 (1.34 - 4.42) | 0.0039 | 0.66 (0.54 - 0.77) | 0.25 (0.08 - 0.46) | 0.15 (0.06 - 0.25) | 0.94 (0.93 - 0.96) |
| CDKN1A | 1.82 (1.19 - 2.75) | 0.0066 | 0.63 (0.48 - 0.76) | 0.33 (0.12 - 0.5) | 0.19 (0.08 - 0.26) | 0.95 (0.93 - 0.96) |
| FURIN | 0.25 (0.09 - 0.70) | 0.0083 | 0.67 (0.54 - 0.79) | 0.29 (0.08 - 0.5) | 0.17 (0.06 - 0.26) | 0.95 (0.93 - 0.96) |
| TNFSF13 | 4.17 (1.43 - 12.15) | 0.0093 | 0.64 (0.54 - 0.75) | 0.25 (0.08 - 0.46) | 0.15 (0.06 - 0.25) | 0.94 (0.93 - 0.96) |
| GAL | 0.16 (0.04 - 0.65) | 0.011 | 0.63 (0.51 - 0.74) | 0.25 (0.08 - 0.46) | 0.15 (0.06 - 0.25) | 0.94 (0.93 - 0.96) |
| TNFSF10 | 0.24 (0.08 - 0.72) | 0.011 | 0.67 (0.55 - 0.77) | 0.17 (0.04 - 0.38) | 0.11 (0.03 - 0.21) | 0.94 (0.93 - 0.95) |
| CPE | 0.31 (0.12 - 0.77) | 0.012 | 0.66 (0.54 - 0.77) | 0.25 (0.08 - 0.46) | 0.15 (0.06 - 0.25) | 0.94 (0.93 - 0.96) |
| WFDC2 | 3.02 (1.27 - 7.34) | 0.012 | 0.64 (0.53 - 0.75) | 0.21 (0.04 - 0.42) | 0.13 (0.03 - 0.23) | 0.94 (0.93 - 0.96) |
| PPY | 0.74 (0.59 - 0.94) | 0.014 | 0.67 (0.56 - 0.78) | 0.21 (0.04 - 0.46) | 0.13 (0.03 - 0.25) | 0.94 (0.93 - 0.96) |
| METAP2 | 1.76 (1.12 - 2.77) | 0.015 | 0.6 (0.44 - 0.74) | 0.29 (0.12 - 0.5) | 0.17 (0.08 - 0.26) | 0.95 (0.93 - 0.96) |
| XPNPEP2 | 0.70 (0.50 - 0.93) | 0.018 | 0.6 (0.49 - 0.7) | 0.17 (0.04 - 0.33) | 0.11 (0.03 - 0.19) | 0.94 (0.93 - 0.95) |
| NECTIN4 | 2.71 (1.19 - 6.04) | 0.018 | 0.61 (0.5 - 0.72) | 0.21 (0.04 - 0.38) | 0.13 (0.03 - 0.21) | 0.94 (0.93 - 0.95) |
| Creatinine | 0.29 (0.10 - 0.86) | 0.024 | 0.62 (0.49 - 0.73) | 0.25 (0.08 - 0.42) | 0.18 (0.07 - 0.27) | 0.93 (0.92 - 0.95) |
| KLK8 | 0.34 (0.13 - 0.88) | 0.026 | 0.61 (0.47 - 0.74) | 0.35 (0.08 - 0.54) | 0.2 (0.06 - 0.28) | 0.95 (0.93 - 0.96) |
| CD207 | 0.35 (0.13 - 0.89) | 0.028 | 0.62 (0.51 - 0.72) | 0.12 (0 - 0.29) | 0.08 (0 - 0.17) | 0.93 (0.93 - 0.95) |
| MIA | 0.24 (0.07 - 0.92) | 0.038 | 0.62 (0.49 - 0.73) | 0.25 (0.08 - 0.42) | 0.15 (0.06 - 0.23) | 0.94 (0.93 - 0.96) |
| GPNUMB | 9.28 (1.10 - 84.13) | 0.040 | 0.63 (0.52 - 0.74) | 0.17 (0.04 - 0.38) | 0.11 (0.03 - 0.21) | 0.94 (0.93 - 0.95) |
| ERBB2 | 3.21 (1.06 - 9.50) | 0.040 | 0.56 (0.41 - 0.71) | 0.33 (0.17 - 0.54) | 0.19 (0.11 - 0.28) | 0.95 (0.94 - 0.96) |
| DLL1 | 3.06 (1.05 - 9.11) | 0.041 | 0.58 (0.45 - 0.7) | 0.25 (0.08 - 0.46) | 0.15 (0.06 - 0.25) | 0.94 (0.93 - 0.96) |
| TNFRSF19 | 0.45 (0.20 - 0.98) | 0.044 | 0.65 (0.51 - 0.77) | 0.33 (0.08 - 0.54) | 0.19 (0.06 - 0.28) | 0.95 (0.93 - 0.96) |
| AR | 1.47 (1.00 - 2.09) | 0.053 | 0.63 (0.51 - 0.74) | 0.21 (0.04 - 0.38) | 0.13 (0.03 - 0.21) | 0.94 (0.93 - 0.95) |
| NT5E | 1.76 (0.99 - 3.08) | 0.053 | 0.63 (0.51 - 0.74) | 0.17 (0.04 - 0.46) | 0.11 (0.03 - 0.25) | 0.94 (0.93 - 0.96) |
| THBS2 | 1.60 (0.98 - 2.69) | 0.060 | 0.62 (0.48 - 0.75) | 0.25 (0.08 - 0.46) | 0.15 (0.06 - 0.25) | 0.94 (0.93 - 0.96) |
| ADAM8 | 0.42 (0.16 - 1.04) | 0.060 | 0.6 (0.48 - 0.73) | 0.25 (0.08 - 0.46) | 0.15 (0.06 - 0.25) | 0.94 (0.93 - 0.96) |
| GZMB | 0.57 (0.30 - 1.06) | 0.077 | 0.61 (0.48 - 0.74) | 0.25 (0.08 - 0.5) | 0.15 (0.06 - 0.26) | 0.94 (0.93 - 0.96) |
| IGF1R | 0.37 (0.12 - 1.12) | 0.078 | 0.64 (0.52 - 0.77) | 0.29 (0.08 - 0.5) | 0.17 (0.06 - 0.26) | 0.95 (0.93 - 0.96) |
| PKM | 1.12 (0.98 - 1.30) | 0.10 | 0.57 (0.43 - 0.72) | 0.12 (0 - 0.46) | 0.1 (0 - 0.29) | 0.92 (0.91 - 0.95) |
| KLK14 | 0.53 (0.25 - 1.16) | 0.11 | 0.59 (0.48 - 0.7) | 0.12 (0 - 0.29) | 0.08 (0 - 0.17) | 0.93 (0.93 - 0.95) |
| ERBB3 | 0.32 (0.08 - 1.32) | 0.11 | 0.59 (0.46 - 0.72) | 0.25 (0.08 - 0.42) | 0.15 (0.06 - 0.23) | 0.94 (0.93 - 0.96) |
| CRNN | 0.67 (0.40 - 1.12) | 0.13 | 0.64 (0.53 - 0.75) | 0.21 (0.04 - 0.42) | 0.13 (0.03 - 0.23) | 0.94 (0.93 - 0.96) |
| FOLR3 | 0.85 (0.64 - 1.05) | 0.14 | 0.57 (0.47 - 0.68) | 0.12 (0 - 0.29) | 0.08 (0 - 0.17) | 0.93 (0.93 - 0.95) |
| CXCL13 | 0.59 (0.28 - 1.18) | 0.14 | 0.58 (0.43 - 0.71) | 0.29 (0.12 - 0.5) | 0.17 (0.08 - 0.26) | 0.95 (0.93 - 0.962) |
| SEZ6L | 0.39 (0.12 - 1.39) | 0.15 | 0.63 (0.5 - 0.75) | 0.29 (0.04 - 0.46) | 0.17 (0.03 - 0.25) | 0.95 (0.93 - 0.96) |
| KLK13 | 0.67 (0.38 - 1.17) | 0.16 | 0.58 (0.45 - 0.72) | 0.25 (0.08 - 0.46) | 0.15 (0.06 - 0.25) | 0.94 (0.93 - 0.96) |
| S100A11 | 0.57 (0.23 - 1.24) | 0.16 | 0.54 (0.42 - 0.66) | 0.21 (0.08 - 0.38) | 0.13 (0.06 - 0.21) | 0.94 (0.93 - 0.95) |
| ITGB5 | 2.09 (0.72 - 6.18) | 0.18 | 0.46 (0.33 - 0.59) | 0.12 (0 - 0.25) | 0.08 (0 - 0.15) | 0.93 (0.93 - 0.94) |
| IL6 | 1.18 (0.91 - 1.46) | 0.18 | 0.61 (0.5 - 0.72) | 0.21 (0.04 - 0.38) | 0.13 (0.03 - 0.21) | 0.94 (0.93 - 0.95) |
| MICA/B | 1.24 (0.91 - 1.88) | 0.19 | 0.62 (0.49 - 0.74) | 0.25 (0.08 - 0.46) | 0.15 (0.06 - 0.25) | 0.94 (0.93 - 0.96) |
| CXCL17 | 1.40 (0.84 - 2.30) | 0.19 | 0.55 (0.46 - 0.65) | 0.08 (0 - 0.21) | 0.06 (0 - 0.13) | 0.93 (0.93 - 0.94) |
| CD70 | 0.62 (0.30 - 1.27) | 0.20 | 0.58 (0.46 - 0.69) | 0.12 (0 - 0.29) | 0.08 (0 - 0.17) | 0.93 (0.93 - 0.95) |
| CD160 | 0.63 (0.30 - 1.28) | 0.20 | 0.57 (0.43 - 0.7) | 0.25 (0.08 - 0.46) | 0.15 (0.06 - 0.25) | 0.94 (0.93 - 0.96) |
| TLR3 | 0.70 (0.41 - 1.22) | 0.20 | 0.59 (0.44 - 0.72) | 0.17 (0 - 0.38) | 0.11 (0 - 0.21) | 0.94 (0.93 - 0.95) |
| VIM | 0.81 (0.55 - 1.17) | 0.27 | 0.55 (0.44 - 0.66) | 0.12 (0 - 0.29) | 0.08 (0 - 0.17) | 0.93 (0.93 - 0.95) |
| IFNGR1 | 1.99 (0.53 - 6.82) | 0.30 | 0.53 (0.39 - 0.65) | 0.12 (0 - 0.29) | 0.08 (0 - 0.17) | 0.93 (0.93 - 0.95) |
| FGFBP1 | 1.70 (0.59 - 4.53) | 0.32 | 0.54 (0.39 - 0.68) | 0.21 (0.08 - 0.46) | 0.13 (0.06 - 0.25) | 0.94 (0.93 - 0.96) |
| LY9 | 0.64 (0.24 - 1.70) | 0.36 | 0.56 (0.43 - 0.69) | 0.25 (0.08 - 0.46) | 0.15 (0.06 - 0.25) | 0.94 (0.93 - 0.96) |
| FOLR1 | 0.68 (0.28 - 1.65) | 0.39 | 0.59 (0.46 - 0.7) | 0.04 (0 - 0.33) | 0.03 (0 - 0.19) | 0.93 (0.93 - 0.95) |
| MSLN | 1.25 (0.74 - 2.05) | 0.40 | 0.58 (0.49 - 0.66) | 0.04 (0 - 0.17) | 0.03 (0 - 0.11) | 0.93 (0.93 - 0.94) |
| PODXL | 2.39 (0.31 - 18.29) | 0.40 | 0.53 (0.4 - 0.67) | 0.21 (0.04 - 0.42) | 0.13 (0.03 - 0.23) | 0.94 (0.93 - 0.96) |
| GZMH | 1.23 (0.74 - 1.95) | 0.41 | 0.47 (0.34 - 0.6) | 0.04 (0 - 0.17) | 0.03 (0 - 0.11) | 0.93 (0.93 - 0.94) |
| S100A4 | 0.81 (0.46 - 1.34) | 0.42 | 0.57 (0.42 - 0.7) | 0.33 (0.17 - 0.5) | 0.19 (0.11 - 0.27) | 0.95 (0.94 - 0.96) |
| ERBB4 | 0.64 (0.20 - 2.08) | 0.45 | 0.57 (0.43 - 0.69) | 0.12 (0 - 0.38) | 0.08 (0 - 0.21) | 0.93 (0.93 - 0.95) |
| TGFB2 | 0.71 (0.28 - 1.75) | 0.46 | 0.59 (0.45 - 0.71) | 0.12 (0 - 0.29) | 0.08 (0 - 0.17) | 0.93 (0.93 - 0.95) |
| SMAD5 | 0.57 (0.15 - 2.95) | 0.46 | 0.61 (0.52 - 0.71) | 0.04 (0 - 0.12) | 0.03 (0 - 0.08) | 0.93 (0.93 - 0.94) |
| VEGFA | 0.78 (0.38 - 1.57) | 0.49 | 0.52 (0.39 - 0.65) | 0.17 (0.04 - 0.33) | 0.11 (0.03 - 0.19) | 0.94 (0.93 - 0.95) |
| CD48 | 0.66 (0.19 - 2.29) | 0.50 | 0.53 (0.4 - 0.66) | 0.21 (0.04 - 0.38) | 0.13 (0.03 - 0.21) | 0.94 (0.93 - 0.95) |
| RSPO3 | 1.16 (0.67 - 1.86) | 0.57 | 0.49 (0.38 - 0.59) | 0 (0 - 0) | 0 (0 - 0) | 0.93 (0.93 - 0.93) |
| CCN4 | 0.82 (0.38 - 1.64) | 0.59 | 0.55 (0.42 - 0.68) | 0.17 (0.04 - 0.33) | 0.11 (0.03 - 0.19) | 0.94 (0.93 - 0.95) |
| IL6ST | 0.87 (0.60 - 2.13) | 0.59 | 0.66 (0.52 - 0.8) | 0.38 (0.21 - 0.58) | 0.21 (0.13 - 0.3) | 0.95 (0.94 - 0.97) |
| LYN | 1.22 (0.39 - 2.70) | 0.69 | 0.53 (0.38 - 0.67) | 0.25 (0.08 - 0.42) | 0.15 (0.06 - 0.23) | 0.94 (0.93 - 0.96) |
| SCAMP3 | 0.94 (0.68 - 1.26) | 0.70 | 0.5 (0.38 - 0.62) | 0 (0 - 0) | 0 (0 - 0) | 0.93 (0.93 - 0.93) |
| CCN1 | 1.11 (0.62 - 2.02) | 0.72 | 0.52 (0.4 - 0.63) | 0 (0 - 0) | 0 (0 - 0) | 0.93 (0.93 - 0.93) |
| SPARC | 0.61 (0.05 - 9.54) | 0.72 | 0.48 (0.37 - 0.59) | 0 (0 - 0) | 0 (0 - 0) | 0.93 (0.93 - 0.93) |
| TNFRSF4 | 1.13 (0.55 - 2.19) | 0.73 | 0.5 (0.37 - 0.64) | 0.17 (0.04 - 0.38) | 0.11 (0.03 - 0.21) | 0.94 (0.93 - 0.95) |
| ABL1 | 0.92 (0.55 - 1.46) | 0.75 | 0.48 (0.34 - 0.6) | 0.08 (0 - 0.21) | 0.06 (0 - 0.13) | 0.93 (0.93 - 0.94) |
| CEACAM1 | 0.83 (0.34 - 4.06) | 0.75 | 0.54 (0.42 - 0.65) | 0.17 (0 - 0.29) | 0.11 (0 - 0.17) | 0.94 (0.93 - 0.95) |
| FADD | 1.07 (0.65 - 1.64) | 0.79 | 0.55 (0.42 - 0.66) | 0.08 (0 - 0.25) | 0.06 (0 - 0.15) | 0.93 (0.93 - 0.94) |
| ADAMTS15 | 1.09 (0.58 - 2.06) | 0.79 | 0.51 (0.39 - 0.64) | 0.12 (0 - 0.25) | 0.08 (0 - 0.15) | 0.93 (0.93 - 0.943) |
| CD27 | 1.09 (0.45 - 2.58) | 0.84 | 0.52 (0.39 - 0.65) | 0.12 (0 - 0.29) | 0.08 (0 - 0.17) | 0.93 (0.93 - 0.95) |
| HGF | 1.05 (0.47 - 2.22) | 0.91 | 0.55 (0.41 - 0.7) | 0.25 (0.08 - 0.42) | 0.15 (0.06 - 0.23) | 0.94 (0.93 - 0.96) |
| KLK11 | 0.96 (0.40 - 2.23) | 0.92 | 0.49 (0.39 - 0.61) | 0.08 (0 - 0.25) | 0.06 (0 - 0.15) | 0.93 (0.93 - 0.94) |
| ANXA1 | 0.98 (0.63 - 1.51) | 0.94 | 0.51 (0.39 - 0.63) | 0.04 (0 - 0.17) | 0.03 (0 - 0.11) | 0.93 (0.93 - 0.94) |
| FLT4 | 0.95 (0.18 - 5.70) | 0.96 | 0.52 (0.4 - 0.65) | 0.08 (0 - 0.21) | 0.06 (0 - 0.13) | 0.93 (0.93 - 0.94) |
| GPC1 | 1.00 (0.37 - 2.75) | 1.00 | 0.54 (0.41 - 0.67) | 0.17 (0.04 - 0.33) | 0.11 (0.03 - 0.19) | 0.94 (0.93 - 0.95) |

**Supplementary Table 4.** ROC AUC, sensitivity (Sens), positive predictive value (PPV) and negative predictive value (NPV) in the validation set of the univariate logistic regression model derived in the discovery set. Markers are ranked according to their respective p values in the discovery set (see Supplementary Table 3). Spec: specificity.

| Marker | ROC AUC | Sens at 90% Spec | PPV at 90% Spec | NPV at 90% Spec |
| --- | --- | --- | --- | --- |
| CA19 - 9 | 0.80 (0.66 - 0.93) | 0.65 (0.48 - 0.87) | 0.49 (0.41 - 0.56) | 0.95 (0.92 - 0.98) |
| KITLG | 0.82 (0.72 - 0.90) | 0.48 (0.22 - 0.70) | 0.41 (0.24 - 0.50) | 0.92 (0.89 - 0.95) |
| CRP | 0.85 (0.76 - 0.92) | 0.59 (0.32 - 0.86) | 0.47 (0.32 - 0.56) | 0.94 (0.90 - 0.98) |
| VWF | 0.74 (0.64 - 0.84) | 0.26 (0.04 - 0.52) | 0.27 (0.06 - 0.43) | 0.89 (0.87 - 0.93) |
| SDC1 | 0.72 (0.59 - 0.83) | 0.30 (0.09 - 0.61) | 0.31 (0.11 - 0.47) | 0.90 (0.87 - 0.94) |
| KDR | 0.77 (0.67 - 0.86) | 0.39 (0.17 - 0.61) | 0.36 (0.20 - 0.47) | 0.91 (0.88 - 0.94) |
| TBIL | 0.67 (0.55 - 0.77) | 0.23 (0.09 - 0.41) | 0.25 (0.12 - 0.38) | 0.89 (0.87 - 0.91) |
| TNFRSF6B | 0.72 (0.60 - 0.83) | 0.39 (0.13 - 0.61) | 0.36 (0.16 - 0.47) | 0.91 (0.88 - 0.94) |
| FASLG | 0.74 (0.64 - 0.84) | 0.26 (0.09 - 0.52) | 0.27 (0.11 - 0.43) | 0.89 (0.87 - 0.93) |
| ESM1 | 0.73 (0.62 - 0.82) | 0.30 (0.13 - 0.52) | 0.31 (0.16 - 0.43) | 0.90 (0.88 - 0.93) |
| EPHA2 | 0.70 (0.56 - 0.82) | 0.39 (0.17 - 0.61) | 0.36 (0.20 - 0.47) | 0.91 (0.88 - 0.94) |
| RET | 0.69 (0.59 - 0.78) | 0.26 (0.09 - 0.43) | 0.27 (0.11 - 0.39) | 0.89 (0.87 - 0.92) |
| TGFA | 0.71 (0.59 - 0.81) | 0.26 (0.09 - 0.48) | 0.27 (0.11 - 0.41) | 0.89 (0.87 - 0.92) |
| MUC16 | 0.71 (0.58 - 0.84) | 0.48 (0.22 - 0.70) | 0.41 (0.24 - 0.50) | 0.92 (0.89 - 0.95) |
| ITGAV | 0.77 (0.67 - 0.85) | 0.39 (0.13 - 0.57) | 0.36 (0.16 - 0.45) | 0.91 (0.88 - 0.94) |
| EGF | 0.74 (0.61 - 0.85) | 0.52 (0.13 - 0.70) | 0.43 (0.16 - 0.50) | 0.93 (0.88 - 0.95) |
| ICOSLG | 0.69 (0.59 - 0.79) | 0.30 (0.09 - 0.48) | 0.31 (0.11 - 0.41) | 0.90 (0.87 - 0.92) |
| CEACAM5 | 0.72 (0.60 - 0.83) | 0.35 (0.13 - 0.52) | 0.33 (0.16 - 0.43) | 0.91 (0.88 - 0.93) |
| CTSV | 0.74 (0.63 - 0.84) | 0.26 (0.09 - 0.57) | 0.27 (0.11 - 0.45) | 0.89 (0.87 - 0.94) |
| TFPI2 | 0.67 (0.54 - 0.78) | 0.22 (0.04 - 0.48) | 0.24 (0.06 - 0.41) | 0.89 (0.87 - 0.92) |
| WIF1 | 0.63 (0.51 - 0.74) | 0.17 (0.00 - 0.39) | 0.20 (0.00 - 0.36) | 0.88 (0.86 - 0.91) |
| TXLNA | 0.64 (0.51 - 0.75) | 0.22 (0.09 - 0.43) | 0.24 (0.11 - 0.39) | 0.89 (0.87 - 0.92) |
| LYPD3 | 0.67 (0.54 - 0.79) | 0.22 (0.04 - 0.43) | 0.24 (0.06 - 0.39) | 0.89 (0.87 - 0.92) |
| CA9 | 0.73 (0.64 - 0.82) | 0.17 (0.04 - 0.35) | 0.20 (0.06 - 0.33) | 0.88 (0.87 - 0.91) |
| FCRLB | 0.69 (0.59 - 0.79) | 0.26 (0.04 - 0.43) | 0.27 (0.06 - 0.39) | 0.89 (0.87 - 0.92) |
| TCL1A | 0.58 (0.45 - 0.70) | 0.17 (0.04 - 0.39) | 0.20 (0.06 - 0.36) | 0.88 (0.87 - 0.91) |
| MDK | 0.70 (0.59 - 0.81) | 0.35 (0.17 - 0.57) | 0.33 (0.20 - 0.45) | 0.91 (0.88 - 0.94) |
| CDKN1A | 0.62 (0.49 - 0.74) | 0.13 (0.00 - 0.43) | 0.16 (0.00 - 0.39) | 0.88 (0.86 - 0.92) |
| FURIN | 0.64 (0.51 - 0.77) | 0.22 (0.04 - 0.48) | 0.24 (0.06 - 0.41) | 0.89 (0.87 - 0.92) |
| TNFSF13 | 0.64 (0.52 - 0.76) | 0.22 (0.04 - 0.43) | 0.24 (0.06 - 0.39) | 0.89 (0.87 - 0.92) |
| GAL | 0.61 (0.47 - 0.72) | 0.17 (0.04 - 0.39) | 0.20 (0.06 - 0.36) | 0.88 (0.87 - 0.91) |
| TNFSF10 | 0.66 (0.54 - 0.77) | 0.22 (0.04 - 0.39) | 0.24 (0.06 - 0.36) | 0.89 (0.87 - 0.91) |
| CPE | 0.64 (0.50 - 0.76) | 0.22 (0.04 - 0.43) | 0.24 (0.06 - 0.39) | 0.89 (0.87 - 0.92) |
| WFDC2 | 0.75 (0.62 - 0.86) | 0.30 (0.09 - 0.61) | 0.31 (0.11 - 0.47) | 0.90 (0.87 - 0.94) |
| PPY | 0.62 (0.51 - 0.73) | 0.17 (0.04 - 0.35) | 0.20 (0.06 - 0.33) | 0.88 (0.87 - 0.91) |
| METAP2 | 0.62 (0.47 - 0.75) | 0.26 (0.09 - 0.48) | 0.27 (0.11 - 0.41) | 0.89 (0.87 - 0.92) |
| XPNPEP2 | 0.57 (0.45 - 0.69) | 0.09 (0.00 - 0.30) | 0.11 (0.00 - 0.31) | 0.87 (0.86 - 0.90) |
| NECTIN4 | 0.62 (0.49 - 0.75) | 0.26 (0.04 - 0.48) | 0.27 (0.06 - 0.41) | 0.89 (0.87 - 0.92) |
| Creatinine | 0.44 (0.29 - 0.58) | 0.09 (0.00 - 0.27) | 0.12 (0.00 - 0.29) | 0.87 (0.86 - 0.89) |
| KLK8 | 0.65 (0.52 - 0.78) | 0.26 (0.04 - 0.48) | 0.27 (0.06 - 0.41) | 0.89 (0.87 - 0.92) |
| CD207 | 0.51 (0.38 - 0.64) | 0.04 (0.00 - 0.17) | 0.06 (0.00 - 0.20) | 0.87 (0.86 - 0.88) |
| MIA | 0.51 (0.40 - 0.63) | 0.04 (0.00 - 0.17) | 0.06 (0.00 - 0.20) | 0.87 (0.86 - 0.88) |
| GNPMB | 0.70 (0.58 - 0.81) | 0.30 (0.09 - 0.52) | 0.31 (0.11 - 0.43) | 0.90 (0.87 - 0.93) |
| ERBB2 | 0.47 (0.34 - 0.61) | 0.13 (0.00 - 0.30) | 0.16 (0.00 - 0.31) | 0.88 (0.86 - 0.90) |
| DLL1 | 0.65 (0.52 - 0.78) | 0.26 (0.04 - 0.52) | 0.27 (0.06 - 0.43) | 0.89 (0.87 - 0.93) |
| TNFRSF19 | 0.52 (0.39 - 0.65) | 0.13 (0.00 - 0.30) | 0.16 (0.00 - 0.31) | 0.88 (0.86 - 0.90) |
| AR | 0.70 (0.57 - 0.81) | 0.22 (0.04 - 0.52) | 0.24 (0.06 - 0.43) | 0.89 (0.87 - 0.93) |
| NT5E | 0.68 (0.55 - 0.80) | 0.22 (0.04 - 0.48) | 0.24 (0.06 - 0.41) | 0.89 (0.87 - 0.92) |
| THBS2 | 0.73 (0.61 - 0.84) | 0.43 (0.22 - 0.65) | 0.39 (0.24 - 0.49) | 0.92 (0.89 - 0.95) |
| ADAM8 | 0.50 (0.38 - 0.62) | 0.09 (0.00 - 0.26) | 0.11 (0.00 - 0.27) | 0.87 (0.86 - 0.89) |
| GZMB | 0.60 (0.47 - 0.73) | 0.22 (0.09 - 0.39) | 0.24 (0.11 - 0.36) | 0.89 (0.87 - 0.91) |
| IGF1R | 0.51 (0.38 - 0.64) | 0.09 (0.00 - 0.22) | 0.11 (0.00 - 0.24) | 0.87 (0.86 - 0.89) |
| PKM | 0.61 (0.46 - 0.75) | 0.23 (0.05 - 0.45) | 0.25 (0.06 - 0.40) | 0.89 (0.86 - 0.92) |
| KLK14 | 0.51 (0.37 - 0.65) | 0.22 (0.04 - 0.39) | 0.24 (0.06 - 0.36) | 0.89 (0.87 - 0.91) |
| ERBB3 | 0.61 (0.50 - 0.72) | 0.13 (0.00 - 0.35) | 0.16 (0.00 - 0.33) | 0.88 (0.86 - 0.91) |
| CRNN | 0.68 (0.57 - 0.78) | 0.17 (0.04 - 0.35) | 0.20 (0.06 - 0.33) | 0.88 (0.87 - 0.91) |
| FOLR3 | 0.54 (0.44 - 0.65) | 0.09 (0.00 - 0.22) | 0.11 (0.00 - 0.24) | 0.87 (0.86 - 0.89) |
| CXCL13 | 0.44 (0.31 - 0.58) | 0.00 (0.00 - 0.13) | 0.00 (0.00 - 0.16) | 0.86 (0.86 - 0.88) |
| SEZ6L | 0.61 (0.48 - 0.73) | 0.17 (0.04 - 0.35) | 0.20 (0.06 - 0.33) | 0.88 (0.87 - 0.91) |
| KLK13 | 0.63 (0.51 - 0.74) | 0.09 (0.00 - 0.26) | 0.11 (0.00 - 0.27) | 0.87 (0.86 - 0.89) |
| S100A11 | 0.58 (0.45 - 0.70) | 0.17 (0.00 - 0.35) | 0.20 (0.00 - 0.33) | 0.88 (0.86 - 0.91) |
| ITGB5 | 0.50 (0.36 - 0.62) | 0.00 (0.00 - 0.13) | 0.00 (0.00 - 0.16) | 0.86 (0.86 - 0.88) |
| IL6 | 0.69 (0.58 - 0.80) | 0.13 (0.00 - 0.39) | 0.16 (0.00 - 0.36) | 0.88 (0.86 - 0.91) |
| MICA/B | 0.58 (0.45 - 0.69) | 0.13 (0.00 - 0.30) | 0.16 (0.00 - 0.31) | 0.88 (0.86 - 0.90) |
| CXCL17 | 0.52 (0.41 - 0.64) | 0.04 (0.00 - 0.17) | 0.06 (0.00 - 0.20) | 0.87 (0.86 - 0.88) |
| CD70 | 0.49 (0.35 - 0.61) | 0.09 (0.00 - 0.26) | 0.11 (0.00 - 0.27) | 0.87 (0.86 - 0.89) |
| CD160 | 0.51 (0.36 - 0.65) | 0.09 (0.00 - 0.26) | 0.11 (0.00 - 0.27) | 0.87 (0.86 - 0.89) |
| TLR3 | 0.68 (0.54 - 0.80) | 0.43 (0.17 - 0.65) | 0.39 (0.20 - 0.49) | 0.92 (0.88 - 0.95) |
| VIM | 0.49 (0.36 - 0.62) | 0.13 (0.00 - 0.26) | 0.16 (0.00 - 0.27) | 0.88 (0.86 - 0.89) |
| IFNGR1 | 0.54 (0.40 - 0.69) | 0.13 (0.00 - 0.35) | 0.16 (0.00 - 0.33) | 0.88 (0.86 - 0.91) |
| FGFBP1 | 0.62 (0.49 - 0.75) | 0.22 (0.09 - 0.43) | 0.24 (0.11 - 0.39) | 0.89 (0.87 - 0.92) |
| LY9 | 0.49 (0.36 - 0.62) | 0.04 (0.00 - 0.17) | 0.06 (0.00 - 0.20) | 0.87 (0.86 - 0.88) |
| FOLR1 | 0.51 (0.37 - 0.63) | 0.09 (0.00 - 0.22) | 0.11 (0.00 - 0.24) | 0.87 (0.86 - 0.89) |
| MSLN | 0.67 (0.56 - 0.79) | 0.35 (0.00 - 0.52) | 0.33 (0.00 - 0.43) | 0.91 (0.86 - 0.93) |
| PODXL | 0.47 (0.34 - 0.61) | 0.09 (0.00 - 0.26) | 0.11 (0.00 - 0.27) | 0.87 (0.86 - 0.89) |
| GZMH | 0.47 (0.33 - 0.60) | 0.13 (0.00 - 0.30) | 0.16 (0.00 - 0.31) | 0.88 (0.86 - 0.90) |
| S100A4 | 0.54 (0.41 - 0.66) | 0.13 (0.00 - 0.26) | 0.16 (0.00 - 0.27) | 0.88 (0.86 - 0.89) |
| ERBB4 | 0.48 (0.34 - 0.60) | 0.09 (0.00 - 0.22) | 0.11 (0.00 - 0.24) | 0.87 (0.86 - 0.89) |
| TGFBR2 | 0.49 (0.36 - 0.63) | 0.00 (0.00 - 0.17) | 0.00 (0.00 - 0.20) | 0.86 (0.86 - 0.88) |
| SMAD5 | 0.62 (0.51 - 0.73) | 0.04 (0.00 - 0.22) | 0.06 (0.00 - 0.24) | 0.87 (0.86 - 0.89) |
| VEGFA | 0.53 (0.40 - 0.65) | 0.09 (0.00 - 0.26) | 0.11 (0.00 - 0.27) | 0.87 (0.86 - 0.89) |
| CD48 | 0.51 (0.39 - 0.64) | 0.04 (0.00 - 0.22) | 0.06 (0.00 - 0.24) | 0.87 (0.86 - 0.89) |
| RSPO3 | 0.48 (0.35 - 0.61) | 0.09 (0.00 - 0.26) | 0.11 (0.00 - 0.27) | 0.87 (0.86 - 0.89) |
| CCN4 | 0.62 (0.49 - 0.74) | 0.22 (0.04 - 0.43) | 0.24 (0.06 - 0.39) | 0.89 (0.87 - 0.92) |
| IL6ST | 0.68 (0.52 - 0.81) | 0.39 (0.22 - 0.61) | 0.36 (0.24 - 0.47) | 0.91 (0.89 - 0.94) |
| LYN | 0.52 (0.39 - 0.65) | 0.22 (0.04 - 0.39) | 0.24 (0.06 - 0.36) | 0.89 (0.87 - 0.91) |
| SCAMP3 | 0.49 (0.38 - 0.61) | 0.00 (0.00 - 0.17) | 0.00 (0.00 - 0.20) | 0.86 (0.86 - 0.88) |
| CCN1 | 0.50 (0.40 - 0.60) | 0.00 (0.00 - 0.04) | 0.00 (0.00 - 0.06) | 0.86 (0.86 - 0.87) |
| SPARC | 0.52 (0.40 - 0.64) | 0.04 (0.00 - 0.26) | 0.06 (0.00 - 0.27) | 0.87 (0.86 - 0.89) |
| TNFRSF4 | 0.64 (0.51 - 0.76) | 0.26 (0.00 - 0.48) | 0.27 (0.00 - 0.41) | 0.89 (0.86 - 0.92) |
| ABL1 | 0.51 (0.39 - 0.63) | 0.00 (0.00 - 0.13) | 0.00 (0.00 - 0.16) | 0.86 (0.86 - 0.88) |
| CEACAM1 | 0.54 (0.41 - 0.66) | 0.17 (0.04 - 0.35) | 0.20 (0.06 - 0.33) | 0.88 (0.87 - 0.91) |
| FADD | 0.55 (0.44 - 0.66) | 0.00 (0.00 - 0.13) | 0.00 (0.00 - 0.16) | 0.86 (0.86 - 0.88) |
| ADAMTS15 | 0.52 (0.39 - 0.65) | 0.13 (0.00 - 0.26) | 0.16 (0.00 - 0.27) | 0.88 (0.86 - 0.89) |
| CD27 | 0.61 (0.47 - 0.74) | 0.26 (0.09 - 0.52) | 0.27 (0.11 - 0.43) | 0.89 (0.87 - 0.93) |
| HGF | 0.51 (0.38 - 0.64) | 0.13 (0.00 - 0.30) | 0.16 (0.00 - 0.31) | 0.88 (0.86 - 0.90) |
| KLK11 | 0.59 (0.46 - 0.70) | 0.13 (0.00 - 0.30) | 0.16 (0.00 - 0.31) | 0.88 (0.86 - 0.90) |
| ANXA1 | 0.50 (0.37 - 0.62) | 0.09 (0.00 - 0.22) | 0.11 (0.00 - 0.24) | 0.87 (0.86 - 0.89) |
| FLT4 | 0.50 (0.36 - 0.62) | 0.13 (0.00 - 0.26) | 0.16 (0.00 - 0.27) | 0.88 (0.86 - 0.89) |
| GPC1 | 0.57 (0.42 - 0.71) | 0.26 (0.09 - 0.48) | 0.27 (0.11 - 0.41) | 0.89 (0.87 - 0.92) |

**Supplementary Table 5.**
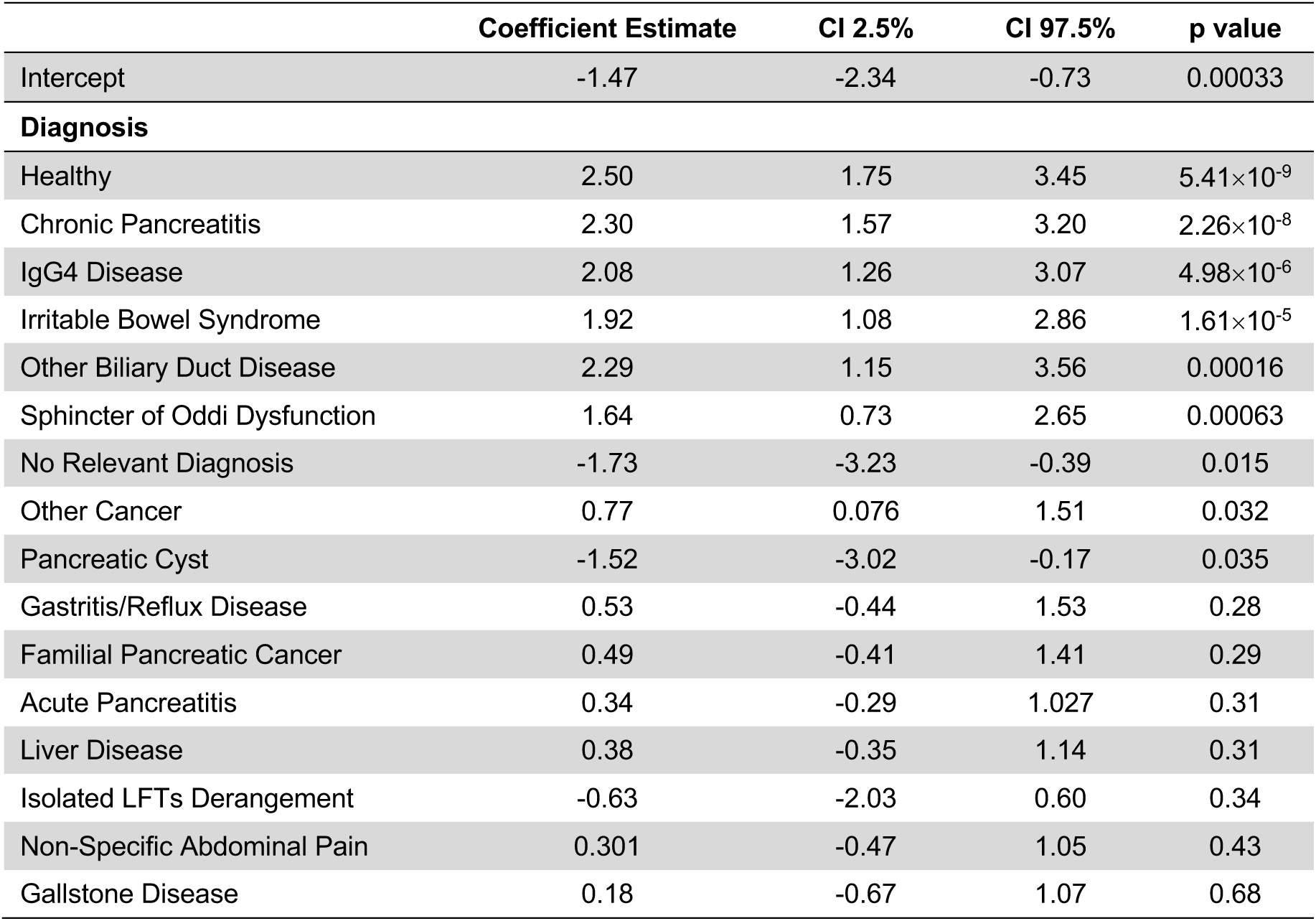
Stack logistic regression model coefficient estimates and respective 95% confidence interval (CI) limits and p values. See statistical analysis sub-section in Methods for the detailed description of the stacking procedure. This model was developed in the discovery set.

**Supplementary Table 6.** Discovery set data and univariate association with PDAC status. Odds ratios (OR), 95% confidence intervals (CI) and p values were calculated according to a logistic regression model with a bias reduction method (see statistical section in Methods).

| Variable |  | Cases | Controls | OR | p value |
| --- | --- | --- | --- | --- | --- |
| Number of samples |  | 24 | 333 | - | - |
| Mean age at sample draw (yr) (range) |  | 70.79 (43.00-91.00) | 58.39(19.00-89.00) | 1.07 (1.03 - 1.11) | 4.68e-05 |
| Mean BMI (kg/m2) (range) |  | 25.46 (19.81-41.35) | 25.41 (15.22-42.19) | 1.01 (0.89-1.12) | 0.87 |
| Gender |  |  |  | 4.98 (2.08 – 13.50) | 0.00023 |
| Male |  | 18 | 121 |  |  |
| Female |  | 6 | 212 |  |  |
| Diabetes |  |  |  |  |  |
| yes |  | 5 | 52 | 1.51 (0.51 – 3.84) | 0.43 |
| no |  | 281 | 19 |  |  |
| Ethnicity |  |  |  |  |  |
| Caucasian |  | 11 | 189 | 1.17 (0.70 – 2.02) | 0.56 |
| Unknown |  | 9 | 99 |  |  |
| Asian |  | 3 | 15 |  |  |
| Other |  | 1 | 13 |  |  |
| Afro/Caribbean |  | 0 | 17 |  |  |

**Supplementary Table 7.** Validation set data and univariate association with PDAC status. Odds ratios (OR), 95% confidence intervals (CI) and p values were calculated according to a logistic regression model with a bias reduction method (see statistical section in Methods).

| Variable |  | Cases | Controls | OR | p value |
| --- | --- | --- | --- | --- | --- |
| Number of Samples |  | 23 | 159 | - | - |
| Mean age at sample draw (yr) (range) |  | 68.61 (53.00 - 83.00) | 57.94 (21.00 - 93.00) | 1.06 (1.02 -1.04) | 0.00071 |
| Mean BMI (kg/m2) (range) |  | 24.18 (12.04 - 31.62) | 25.64 (17.60 - 38.30) | 0.90 (0.78-1.01) | 0.080 |
| Gender |  |  |  | 2.65 (1.11 – 6.58) | 0.028 |
| Male |  | 14 | 58 |  |  |
| Female |  | 9 | 101 |  |  |
| Diabetes |  |  |  |  |  |
| yes |  | 5 | 52 | 1.57 (0.51 – 4.23) | 0.41 |
| no |  | 281 | 19 |  |  |
| Ethnicity |  |  |  |  |  |
| Caucasian |  | 10 | 102 | 2.66 (1.42 – 5.17) | 0.0020 |
| Unknown |  | 12 | 32 |  |  |
| Asian |  | 3 | 15 |  |  |
| Other |  | 1 | 5 |  |  |
| Afro/Caribbean |  | 0 | 5 |  |  |

**Supplementary Table 8.** Type and number of subjects with other cancers in the ADEPTS cohort. See also Figure 1.

| Other Cancer | Number of subjects |
| --- | --- |
| Possible gallbladder cancer | 1 |
| Low grade dysplasia on ampulla of Vater (Incidental finding) | 1 |
| Hilar cholangiocarcinoma, treated Nov 2017 | 1 |
| Low anal moderately differentiated adenocarcinoma | 1 |
| Cholangiocarcinoma, primary sclerosing cholangitis | 1 |
| PNET (insulinoma) | 1 |
| Bowel cancer in 2011 | 1 |
| Prostate cancer | 1 |

**Supplementary Table 9.** Values corresponding to Figure 4. Only ADEPTS samples were considered for this association study. Given that symptoms were not considered in the training of the classifiers, we concatenate ADEPTS samples in the discovery and validation sets to verify the associations of symptoms and PDAC. A univariate logistic regression model with bias correction was used for each symptom to test the association with PDAC.

| Symptoms | Number of subjects |  |  |  | OR (95% CI) | p value |
| --- | --- | --- | --- | --- | --- | --- |
|  | Yes |  | No |  |  |  |
|  | Control | Case | Control | Case |  |  |
| Jaundice | 18 | 22 | 401 | 23 | 20.78 (9.98 - 44.35) | 3.22×10<sup>-15</sup> |
| Weight Loss | 39 | 17 | 380 | 28 | 5.91 (2.96 - 11.62) | 1.44×10<sup>-06</sup> |
| Asymptomatic | 96 | 3 | 323 | 42 | 0.28 (0.07- 0.74) | 0.0077 |
| Reflux | 38 | 0 | 381 | 45 | 0.11 (0.00 - 0.79) | 0.022 |
| Bloating | 31 | 0 | 388 | 45 | 0.14 (0.00 - 0.99) | 0.048 |
| Dyspepsia | 30 | 0 | 389 | 45 | 0.14 (0.00 - 1.03) | 0.054 |
| Abdominal Pain | 198 | 16 | 221 | 29 | 0.62 (0.33 - 1.16) | 0.14 |
| Nausea | 20 | 0 | 399 | 45 | 0.21 (0.00 - 1.60) | 0.17 |
| Vomiting | 19 | 4 | 400 | 41 | 2.23 (0.67 - 6.03) | 0.17 |
| Asymptomatic<br>LFT Derangement | 52 | 8 | 367 | 37 | 1.59 (0.67 - 3.39) | 0.28 |
| Anaemia | 24 | 1 | 395 | 44 | 0.54 (0.06 - 2.18) | 0.44 |
| Back Pain | 1 | 0 | 418 | 45 | 3.07 (0.02 - 58.34) | 0.54 |
| Heartburn | 9 | 0 | 410 | 45 | 0.47 (0.00 - 3.85) | 0.57 |
| Change In Bowel Habit | 58 | 7 | 361 | 38 | 1.20 (0.49 - 2.62) | 0.67 |
| Rectal Bleeding | 10 | 1 | 409 | 44 | 1.31 (0.14 - 5.80) | 0.76 |
| Dysphagia | 16 | 1 | 403 | 44 | 0.82 (0.09 - 3.42) | 0.82 |

**Supplementary Table 10.**
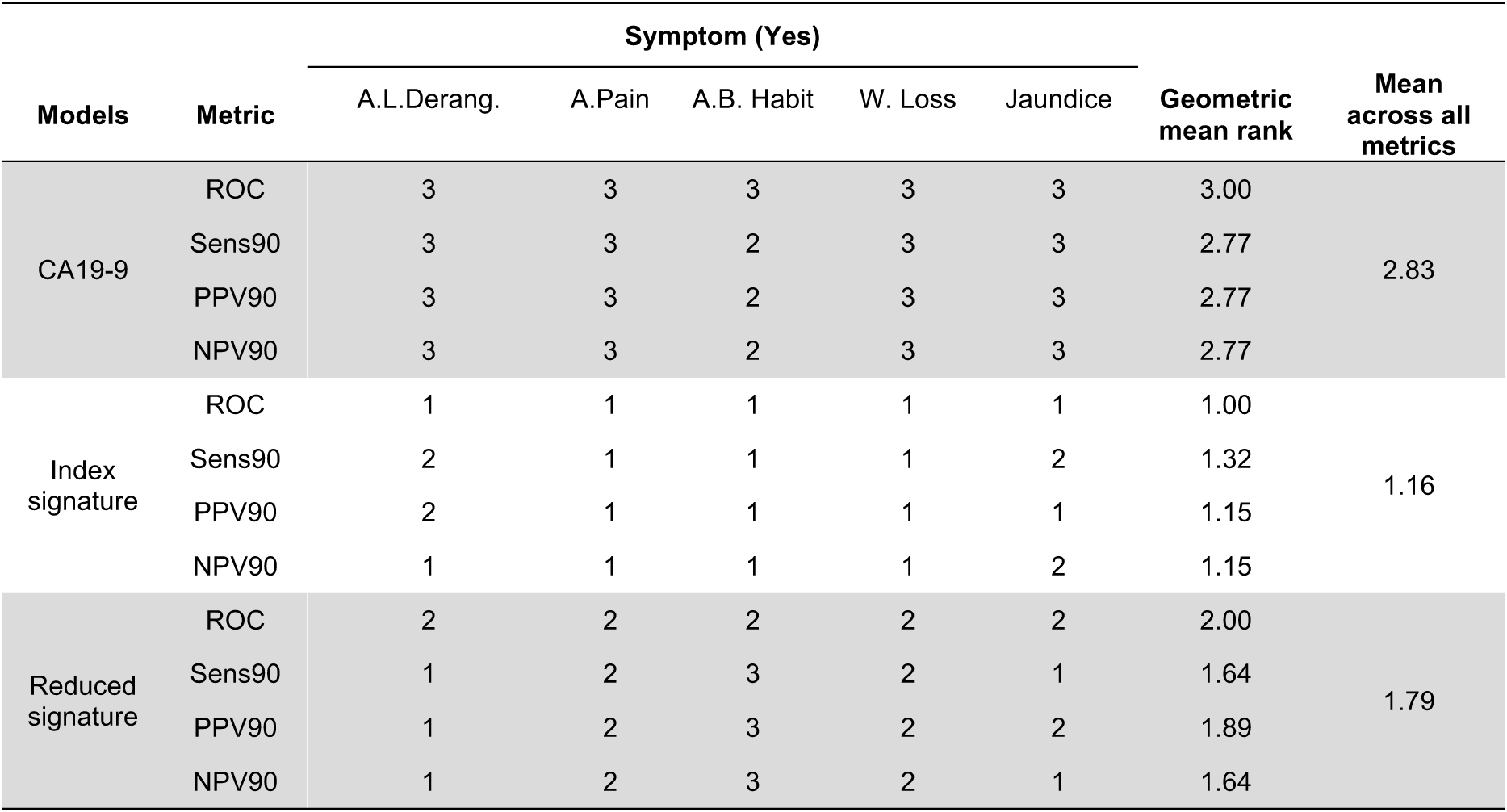
Performance model rank summary for selected models in symptomatic patients. The probability values used to calculate the performance metrics were generated with each model developed in the training set and reported in the main text. Probability values for symptomatic patients belonging to the training set and validation set were concatenated to generate the ROC curves. Only ADEPTS samples had symptoms information. A. L. Derang.: Asymptomatic LFT Derangement. B. Pain: Back Pain. C. B. Habit: Change in Bowel Habit. W. Loss: Weight Loss. Here, only the ranks of the performances are provided. For the respective performance values see Table 2 in the main text.

| Models | Metric | Symptom (Yes) |  |  |  |  | Geometric mean rank | Mean across all metrics |
| --- | --- | --- | --- | --- | --- | --- | --- | --- |
|  |  | A.L.Derang. | A.Pain | A.B. Habit | W. Loss | Jaundice |  |  |
| CA19-9 | ROC | 3 | 3 | 3 | 3 | 3 | 3.00 | 2.83 |
|  | Sens90 | 3 | 3 | 2 | 3 | 3 | 2.77 |  |
|  | PPV90 | 3 | 3 | 2 | 3 | 3 | 2.77 |  |
|  | NPV90 | 3 | 3 | 2 | 3 | 3 | 2.77 |  |
| Index signature | ROC | 1 | 1 | 1 | 1 | 1 | 1.00 | 1.16 |
|  | Sens90 | 2 | 1 | 1 | 1 | 2 | 1.32 |  |
|  | PPV90 | 2 | 1 | 1 | 1 | 1 | 1.15 |  |
|  | NPV90 | 1 | 1 | 1 | 1 | 2 | 1.15 |  |
| Reduced signature | ROC | 2 | 2 | 2 | 2 | 2 | 2.00 | 1.79 |
|  | Sens90 | 1 | 2 | 3 | 2 | 1 | 1.64 |  |
|  | PPV90 | 1 | 2 | 3 | 2 | 2 | 1.89 |  |
|  | NPV90 | 1 | 2 | 3 | 2 | 1 | 1.64 |  |

**Supplementary Table 11.** Pairwise area under receiver operating characteristic curve comparison p-values for the selected models in Table 2 (main text). Only ADEPTS samples had symptoms information. A. L. Derang.: Asymptomatic LFT Derangement. B. Pain: Back Pain. C. B. Habit: Change in Bowel Habit. W. Loss: Weight Loss. For the respective performance values see Table 2 in the main text. Models in rows for each symptom are compared with those in the columns for the same symptom. 10000 bootstraps were constructed to test the significance of the difference in performance being lower.

| Symptom (Yes) |  | Reduced signature | Index signature |
| --- | --- | --- | --- |
| A.L.Derang. | CA19-9 | $3.89 \times 10^{-07}$ | $7.70 \times 10^{-26}$ |
|  | Reduced signature | - | 0.25 |
| A. Pain | CA19-9 | $2.19 \times 10^{-15}$ | $2.83 \times 10^{-93}$ |
| | Reduced signature | - | $1.08 \times 10^{-06}$ |
| A. B. Habit | CA19-9 | 0.0018 | $1.48 \times 10^{-28}$ |
|  | Reduced signature | - | 0.020 |
| W.Loss | CA19-9 | $2.04 \times 10^{-09}$ | $1.38 \times 10^{-14}$ |
|  | Reduced signature |  | 0.018 |
| Jaundice | CA19-9 | 0.013 | $3.34 \times 10^{-07}$ |
|  | Reduced signature | - | 0.069 |

